# School closures and SARS-CoV-2. Evidence from Sweden’s partial school closure

**DOI:** 10.1101/2020.10.13.20211359

**Authors:** Jonas Vlachos, Edvin Hertegård, Helena Svaleryd

## Abstract

To reduce the transmission of SARS-CoV-2 most countries closed schools, despite uncertainty if school closures are an effective containment measure. At the onset of the pandemic, Swedish upper secondary schools moved to online instruction while lower secondary school remained open. This allows for a comparison of parents and teachers differently exposed to open and closed schools, but otherwise facing similar conditions. Leveraging rich Swedish register data, we connect all students and teachers in Sweden to their families and study the impact of moving to online instruction on the incidence of SARS-CoV-2 and COVID-19. We find that among parents, exposure to open rather than closed schools resulted in a small increase in PCR-confirmed infections [OR 1.17; CI95 1.03–1.32]. Among lower secondary teachers the infection rate doubled relative to upper secondary teachers [OR 2.01; CI95 1.52–2.67]. This spilled over to the partners of lower secondary teachers who had a higher infection rate than their upper secondary counterparts [OR 1.29; CI95 1.00–1.67]. When analyzing COVID-19 diagnoses from healthcare visits and the incidence of severe health outcomes, results are similar for teachers but weaker for parents and teachers’ partners. The results for parents indicate that keeping lower secondary schools open had minor consequences for the overall transmission of SARS-CoV-2 in society. The results for teachers suggest that measures to protect teachers could be considered.

## 1. Introduction

In the effort to contain the spread of SARS-CoV-2 most countries closed schools during the ongoing pandemic. An estimated 1.3 billion students in 195 countries were affected by school closures in mid-April 2020 (UNESCO, 2020). These closures are likely to have a negative impact on student learning and well-being, especially for students from disadvantaged backgrounds (Dorn, Hancock, Sarakatsannis & Viruleg, 2020; Guessoum et al., 2020). School closures also affect labor supply, not least among healthcare workers, hence reducing healthcare capacity (Bayham & Fenichel, 2020). While the costs associated with school closures are high, modelling studies question their effectiveness in reducing the transmission of SARS-CoV-2 and direct evidence is largely missing (Viner et al., 2020). The absence of direct evidence is because school closures were usually implemented early, universally, and in close proximity to a raft of non-pharmaceutical interventions (NPIs) that have been documented and modelled to bring about large reductions in the basic reproduction number (Hsiang et al., 2020; Kraemer et al., 2020; Pan et al., 2020; Tian et al., 2020; Maier & Brockmann, 2020; Auger et al., 2020). This renders it difficult, if not impossible, to disentangle the effects of each specific intervention.

Sweden was an exception to the norm of universal school closures. On March 18, 2020, one week after the first reported death from COVID-19, upper secondary schools moved to online instruction while schools for younger students remained open until the end of the school year in mid-June. While other NPIs were also implemented (see supplement), this partial school closure allows for a comparison of individuals and households who were differently exposed to open and closed schools, but otherwise faced similar conditions throughout the period of widespread contagion illustrated in Fig. 1. In this study, we link detailed register data from *Statistics Sweden* on the entire Swedish population to all PCR (Polymerase Chain Reaction) identified cases of SARS-CoV-2 reported to the *Public Health Agency of Sweden* and COVID-19 cases requiring medical treatment reported to the *National Board of Health and Welfare* between the time of school closure to the end of the school year. To study the general impact of school closure on the transmission of the virus, we estimate differences in infection rates between parents exposed to lower and upper secondary students. We further analyze differences in infection rates between lower and upper secondary teachers as well as their partners.

**Fig. 1:**
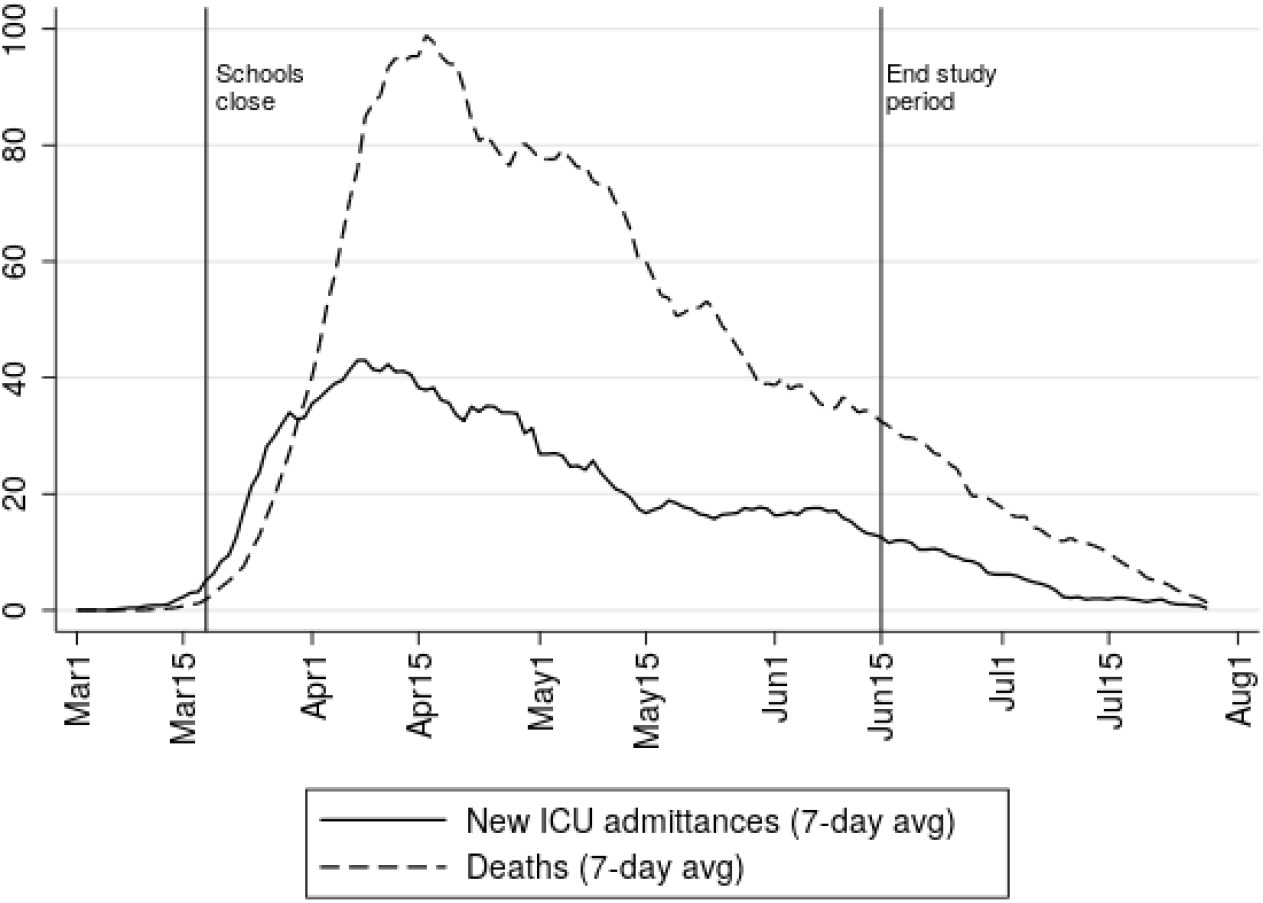
COVID-19 deaths and ICU admissions. 7-day averages of deaths and ICU admissions. Solid vertical lines mark the start of school closure and the end of the period of analysis. Data from the Public Health Agency of Sweden (Public Health Agency of Sweden, 2020a).

For school closures to affect virus transmission, they must affect behavior and contact patterns. The impact of school closures on the transmission of SARS-CoV-2 further depends on how the virus spreads between students, from students to adults, and among adults in school and at home. Current reviews of the evidence suggest that while children and adolescents do get infected, they usually develop mild or no symptoms (ECDC, 2020; Goldstein, Lipsitch & Cevik, 2020). The susceptibility to infection appears to be lower among the young, but there is some uncertainty regarding this as a large number of cases probably go undetected. Children and adolescents with mild or no symptoms may still carry and spread the infection, but the evidence available indicates that infectiousness, just as the severity of symptoms, is increasing in age. Outbreaks have been reported in connection to school openings and overnight summer camps (Stein-Zamir et al., 2020; Szablewski, 2020), but transmission within schools prior to their closure at the onset of the pandemic appears to have been limited (Heavey, Casey, Kelly, Kelly & McDarby, 2020; Macartney et al., 2020). A general caveat concerning the available evidence is that most studies on the susceptibility and infectiousness of children and adolescents have been conducted when schools were closed and other NPIs were in place.

Differences between groups can be attributed to school closures if the groups are behaviorally and biologically similar in all other respects that affect the probability to get infected and tested. Lower secondary school (school years 7–9, typical age 14–16) is compulsory. Attendance to upper secondary school (school years 10–12, typical age 17–19) is close to universal but grade repetition is more common at the upper-secondary level, in particular among students with non-EU background (Swedish National Agency for Education, 2020a). We therefore restrict the main sample to parents without such a background, but also present results for all parents. The main selection concern regards the age of parents and students. Parental characteristics (age, sex, income, occupation, region of origin and of residence) are controlled for, but the susceptibility and infectiousness are likely to increase in student age and general behavior may differ between younger and older students. We therefore focus our attention on parents exposed to students in the final year of lower secondary and first year of upper secondary school. The main concern regarding differences between upper and lower secondary teachers and their partners refers to partner characteristics that are adjusted for. Given these restrictions and adjustments, the estimated differences can plausibly be attributed to the exposure to open and closed schools. The study thus offers credible direct evidence on the impact of school closures on the SARS-CoV-2 pandemic.

Models predict that school closures can be effective if they actually reduce the number of contacts, the basic reproduction number (R0) is below 2, and the attack rate is higher in children than in adults (Jackson, Mangtani, Hawker, Olowokure & Vynnycky, 2014). The basic reproduction number for SARS-CoV-2 is above 2 (C.-C. Lai, Shih, Ko, Tang & Hsueh, 2020) and the attack rate in students is likely to be low relative to adults (ECDC, 2020). The theoretical prior is therefore that the impact of school closures on the transmission of SARS-CoV-2 among parents is low (Viner et al., 2020). For teachers and their partners, a more substantive impact can be expected. Teachers at open schools were not only exposed to students but also to other adults, both at work and during their commute. Upper secondary teachers partly worked from school, but a substantive fraction did their teaching from home (see SI Appendix).

## 2. Results

We estimate differences in infections among parents, teachers, and teachers’ partners who were differently exposed to lower (open) and upper (online) secondary schools using linear probability models (OLS) and logistic regressions (Logit). Descriptive statistics are shown in Table 2. The preferred outcome is PCR-confirmed SARS-CoV-2 which has the highest incidence (7.37 cases/1000 among lower and upper secondary parents and 4.69/1000 among teachers). If we exclude healthcare workers who were targeted for testing, the incidence drops to 4.33/1000 among parents. One potential drawback of this outcome is that unbiased results rely on compared groups having equal propensity to get tested. In particular, it could be that those directly or indirectly exposed to open schools were more prone to get tested which would exaggerate the impact of school closures. The risk of such bias is alleviated by the limited testing capacity that forced testing to be targeted towards those with severe symptoms and care workers throughout most of the relevant period (see SI Appendix). However, we also analyze COVID-19 diagnoses from healthcare visits which is less likely to suffer from bias due to behavioral differences. Healthcare coverage in Sweden is universal and fees for doctor or hospital visits are low, assuring individuals in need will seek care. This is particularly true for hospitalizations since admittance to hospital is determined strictly on medical grounds. As receiving a COVID-19 diagnosis is a less frequent event (3.11/1000 among parents; 2.60/1000 among teachers) these estimations have lower statistical power. Low incidence is an even larger problem for severe cases (hospitalizations or deaths) which has an incidence of 1.43/1000 among parents and 1.59/1000 among teachers. Results for severe cases reported in SI Appendix, Table S1.

**Table 1:**
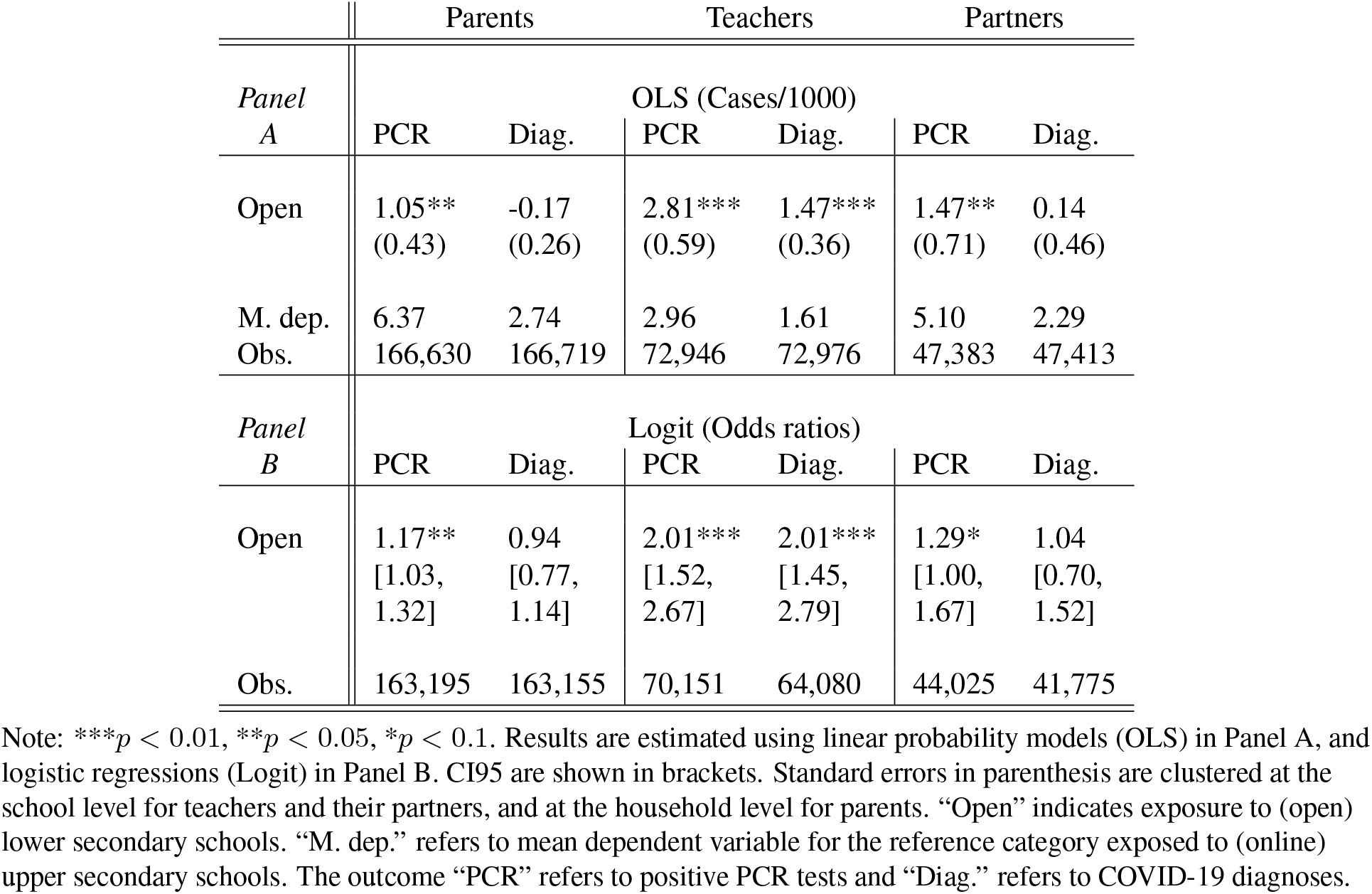
Effect of exposure to open schools on PCR tests and COVID-19 diagnoses

|  | Parents |  | Teachers |  | Partners |  |
| --- | --- | --- | --- | --- | --- | --- |
| Panel<br>A | OLS (Cases/1000) |  |  |  |  |  |
|  | PCR | Diag. | PCR | Diag. | PCR | Diag. |
| Open | 1.05**<br>(0.43) | -0.17<br>(0.26) | 2.81***<br>(0.59) | 1.47***<br>(0.36) | 1.47**<br>(0.71) | 0.14<br>(0.46) |
| M. dep.<br>Obs. | 6.37<br>166,630 | 2.74<br>166,719 | 2.96<br>72,946 | 1.61<br>72,976 | 5.10<br>47,383 | 2.29<br>47,413 |
| Panel<br>B | Logit (Odds ratios) |  |  |  |  |  |
|  | PCR | Diag. | PCR | Diag. | PCR | Diag. |
| Open | 1.17**<br>[1.03,<br>1.32] | 0.94<br>[0.77,<br>1.14] | 2.01***<br>[1.52,<br>2.67] | 2.01***<br>[1.45,<br>2.79] | 1.29*<br>[1.00,<br>1.67] | 1.04<br>[0.70,<br>1.52] |
| Obs. | 163,195 | 163,155 | 70,151 | 64,080 | 44,025 | 41,775 |

Note: \*\*\* $p < 0.01$ , \*\* $p < 0.05$ , \* $p < 0.1$ . Results are estimated using linear probability models (OLS) in Panel A, and logistic regressions (Logit) in Panel B. CI95 are shown in brackets. Standard errors in parenthesis are clustered at the school level for teachers and their partners, and at the household level for parents. “Open” indicates exposure to (open) lower secondary schools. “M. dep.” refers to mean dependent variable for the reference category exposed to (online) upper secondary schools. The outcome “PCR” refers to positive PCR tests and “Diag.” refers to COVID-19 diagnoses.

**Table 2:** Descriptive statistics

|  | Parents school years 7–12 |  |  | Parents school years 9–10 |  |  |
| --- | --- | --- | --- | --- | --- | --- |
|  | Full sample | Lower sec. | Upper sec. | Full sample | Lower sec. | Upper sec. |
| Cases/1000 | 7.37 | 7.20 | 7.56 | 7.57 | 8.00 | 7.15 |
| ... ex health | 4.33 | 4.23 | 4.43 | 4.51 | 4.70 | 4.32 |
| ...pre cut-off | 0.64 | 0.56 | 0.74 | 0.73 | 0.67 | 0.79 |
| Healthcare/1000 | 3.11 | 2.91 | 3.34 | 3.22 | 3.06 | 3.37 |
| Severe cases/1000 | 1.43 | 1.27 | 1.61 | 1.48 | 1.37 | 1.59 |
| # Deaths | 25 | 9 | 16 | 6 | 3 | 3 |
| Age | 50.27<br>(5.89) | 48.89<br>(5.76) | 51.81<br>(5.65) | 50.46<br>(5.69) | 49.98<br>(5.66) | 50.92<br>(5.69) |
| Obs. | 480,291 | 253,538 | 226,753 | 166,630 | 81,598 | 85,032 |

|  | Teachers |  |  | Teachers' partners |  |  |
| --- | --- | --- | --- | --- | --- | --- |
|  | Full sample | Lower sec. | Upper sec. | Full sample | Lower sec. | Upper sec. |
| Cases/1000 | 4.69 | 5.91 | 3.25 | 6.16 | 6.60 | 5.64 |
| ... ex health |  |  |  | 4.21 | 4.99 | 3.26 |
| ...pre cut-off | 0.27 | 0.25 | 0.30 | 0.59 | 0.63 | 0.55 |
| Healthcare/1000 | 2.60 | 3.29 | 1.79 | 2.65 | 2.77 | 2.51 |
| Severe cases/1000 | 1.59 | 2.00 | 1.10 | 1.25 | 1.33 | 1.16 |
| # Deaths | 1 | 1 | 0 | 1 | 1 | 0 |
| Age | 47.84<br>(10.61) | 47.37<br>(10.58) | 48.39<br>(10.62) | 49.17<br>(10.14) | 49.01<br>(10.18) | 49.36<br>(10.08) |
| Obs. | 72,946 | 39,446 | 33,500 | 47,383 | 25,587 | 21,796 |

Note: The table shows descriptive statistics for the three study populations. “Cases/1000” denotes positive PCR tests per 1000 until June 15, 2020. “...ex. health” means that healthcare and care workers are dropped (occupational codes 15, 22, 32, 53). “...pre cut-off” refers to cases before the specified cut-off dates referring to school closures. Cut-off dates are March 25 for teachers, and April 1 for parents and teachers’ partners. “Healthcare/1000” shows open care, inpatient care, and deaths related to COVID-19 per 1000, reported until June 30. “Severe cases/1000” shows only inpatient care and deaths related to COVID-19 per 1000, reported until June 30. The number of deaths shows reported deaths before July 26 among those tested positive until June 15. Standard deviation for age is shown in parenthesis. The number of observations refers to the sample of individuals with a positive or no PCR test during the study period. Individuals with a positive PCR test with an invalid date are excluded.

The data covers the entire relevant Swedish population and contains all reported cases as well as detailed information on covariates (see Materials and Methods and SI Appendix for details). Upper secondary schools moved online on March 18. Allowing for an incubation period from infection to symptoms of about a week (Lauer et al., 2020), the cut-off date is set to March 25 for teachers and April 1 for parents and teachers’ partners. The school year ends during the second week of June and the end date is therefore set to June 15 for PCR tests and June 30 for diagnoses through healthcare contacts.

### Parents

Parental school exposure is defined by the school year that the youngest child in the household attends. In order to attribute estimated differences to school closures, households must be similar in all aspects that affect the likelihood of getting infected or tested, except for their exposure to open and closed schools. By narrowing the comparison to parents with the youngest child in the final year of lower secondary (Year 9) and first year of upper secondary school (Year 10), we reduce the risk of introducing biases due to confounding factors. A potential threat to identification is that student with non-EU migrant background are more likely to repeat grades in upper secondary schools, in particular through preparatory programs (Swedish National Agency for Education, 2020a). Although upper secondary grade repetition occurs also for other groups, the concern is not as severe among families from Sweden, the EU, and the Nordic countries. To avoid selection into grade 10 in upper secondary school, we restrict the population to parents born in Sweden, EU and the Nordics (dropping 16% of the parental population). In SI Appendix, we substantiate these claims by showing balance on covariates predicting the incidence of SARS-CoV-2 for the main sample (SI Appendix, Fig. S2) while balancing tests perform worse when including non-EU migrants (SI Appendix, Fig. S3).

Fig. 2 shows the estimated odds ratios for PCR-confirmed SARS-CoV-2 parents from logistic regressions where we adjust for age, sex, occupation, educational attainment, income, regions of residence and of origin. Results for parents by school years 7–12 show that there is a tendency of a positive age gradient, potentially indicating a higher parental risk of infection when exposed to older children. The most relevant comparison is therefore between school years 9 and 10 (reference category) for which we in Table 1 estimate an odds ratio of 1.17 [CI95 1.03–1.32].

**Fig. 2:**
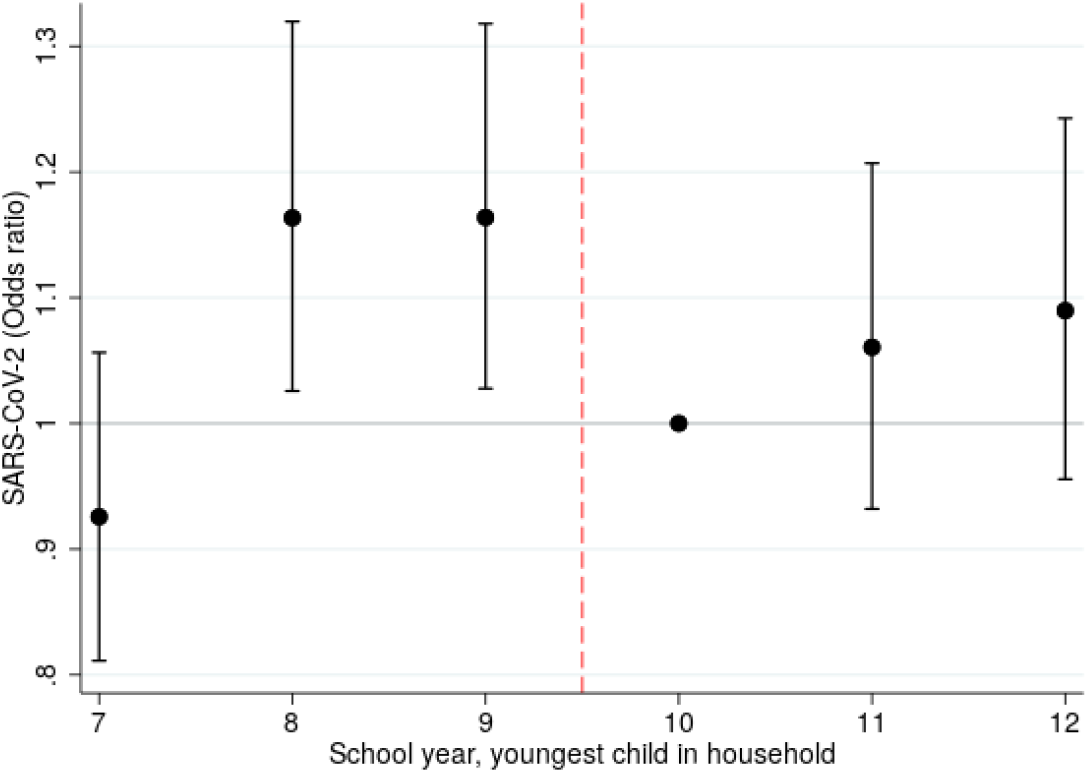
SARS-CoV-2 odds ratios for parents by school year of the youngest child in the household. Odds ratios estimated using logistic regression. The reference category is school year 10 and 95% confidence intervals are indicated.

Corresponding results using OLS are shown in Table 1 which also includes results for COVID-19 diagnoses from healthcare contacts. The estimates indicate that parental exposure to open schools results in 1.05 (se 0.43) additional SARS-CoV-2 cases per 1000 individuals and −0.17 (se 0.26) additional COVID-19 diagnoses per 1000. The odds ratio for COVID-19 diagnoses is 0.94 [CI95 0.77–1.14]. The estimates for COVID-19 diagnoses are thus negative, albeit imprecise and statistically indistinguishable from zero. This indicates that the increase in PCR-confirmed cases does not necessarily translate into similar size effects on the probability to get a COVID-19 diagnoses when visiting a doctor or being admitted to hospital. The same applies to the estimates for severe cases shown in SI Appendix, Table S1 [OR 0.84; CI95 0.64–1.11].

### Teachers

We analyze differences between lower and upper secondary teachers and their partners. Upper secondary teachers constitute a relevant counterfactual to the work situation that lower secondary teachers had been in if their schools had moved to online instruction. The groups are also similar with respect to educational attainment and geographic dispersion. As there may still be differences in the household composition between the groups, we — in addition to the controls used for parents — adjust for the occupation and educational attainment of teachers’ partners, the number of children in separate age groups linked to the household, and whether or not the teacher is single. Table 1 shows that the likelihood of a positive PCR test was twice as high for lower secondary than for upper secondary teachers [OR 2.01; CI95 1.52–2.67]. The table also shows a corresponding OLS estimate of 2.81 additional cases/1000 (se 0.59). An identical estimate is found for COVID-19 diagnoses [OR 2.01; CI95 1.45–2.79], indicating that the PCR results are not due to biased testing. Table S1 in SI Appendix shows an estimate for severe cases of similar magnitude [OR 2.15; CI95 1.41–3.29].

In order to gauge the magnitude of the estimated effects for teachers, Fig. 3 compares the incidence of detected SARS-CoV-2 among teachers with occupations at the three-digit level with at least 1000 employees in ages 25–65 (healthcare workers excluded). Among the 124 compared occupations, upper secondary teachers (3.25/1000) are at the median while lower secondary teachers (5.91/1000) constitute the 7th most affected occupation. Drivers (which includes taxi drivers) are the at the top of the distribution while driving instructors have the same level of infections as lower secondary teachers.

**Fig. 3:**
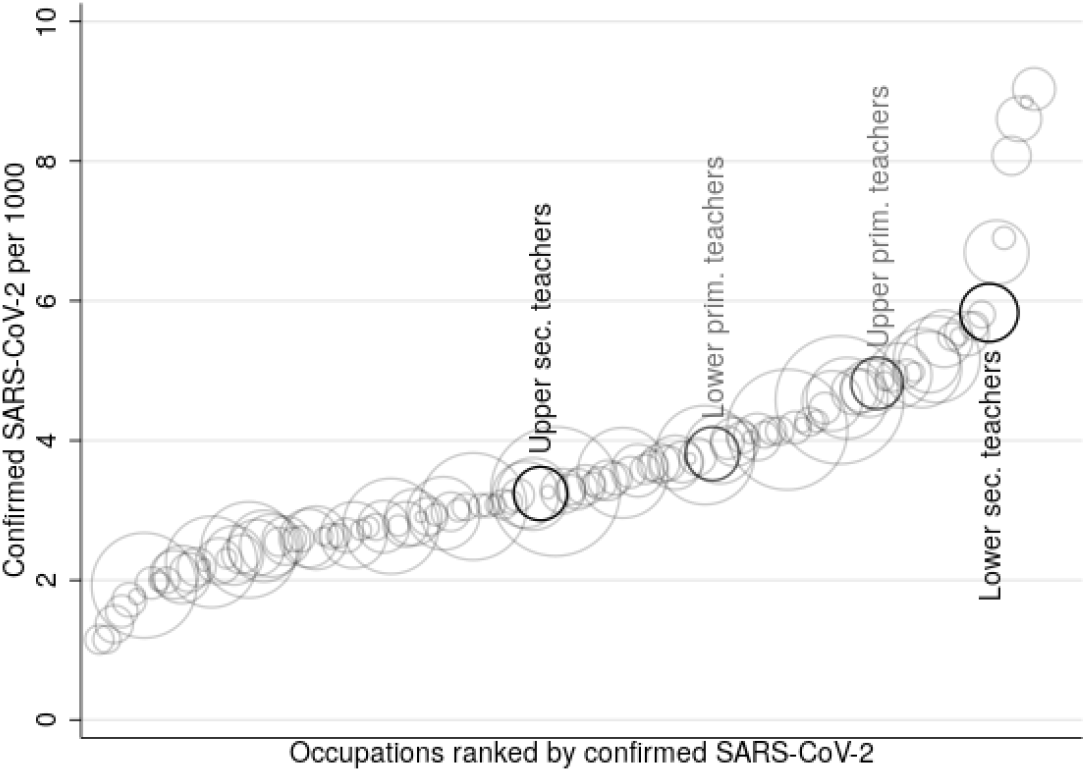
SARS-CoV-2 across occupations. Circle size corresponds to the number of employees in each occupation. Incidence (cases per 1000) of detected SARS-CoV-2 by 3-digit occupational codes (SSYK2012) until June 15, 2020. Ages 25–65, only occupations with at least 1000 employees reported. Values for the upper and lower secondary teachers (as well as lower and upper primary teachers in grey) from the Teacher Register in our sample indicated in black.

A list of all occupations is available in SI Appendix, Table S11. As another comparison, Table 2 shows that the incidence of SARS-CoV-2 is higher among lower secondary teachers (5.91/1000) than that of the parents to the students they teach (4.23/1000, excluding healthcare workers). This is also the case for COVID-19 diagnoses and more severe health outcomes. Note that parents with non-EU background are excluded from these comparisons. The rate of infections is higher than average in this group and when included, the rate among lower secondary parents increases to 5.33 cases/1000 (excluding healthcare).

Fig. 3 also indicates the incidence of detected SARS-CoV-2 among lower primary (school years 1–3; 3.81 cases/1000) and upper primary (years 4–6; 4.82 cases/1000) teachers. These teachers are less specialized and therefore not only meet younger but also fewer students than teachers at the lower secondary level. They may also interact differently with their colleagues. The incidence among these teachers is below lower secondary teachers but above upper secondary teachers, also when controlling for covariates (SI Appendix, Table S2). These results are consistent with a positive risk gradient in student age, but could reflect other differences in the work environment.

### Teachers’ partners

The higher incidence of infections among lower secondary teachers spilled over to their partners who have a higher incidence of positive PCR tests than their upper secondary counterparts [OR 1.29; CI95 1.00–1.67] (Table 1). This is evidence of within household transmission from teachers to their partners. The estimates for teachers and their partners implies a secondary attack rate (SAR) between spouses of 0.52 [CI95 0.05–1.18].^1^ This is well within the bounds of the between-spouse SAR of 0.43 [CI95 0.27–0.6] suggested from contact studies (Madewell, Yang, Longini, Halloran & Dean, 2020). However, the estimates for COVID-19 diagnoses for teachers’ partners are not statistically distinguishable from zero [OR 1.04; CI95 0.70–1.52] and the same applies for severe cases [OR 1.09; CI95 0.62–1.92] (SI Appendix, Table S1). The relatively imprecise estimates for these outcomes also renders them statistically indistinguishable from the estimates for PCR-confirmed SARS-CoV-2.

### Robustness

In the SI Appendix, we provide several robustness tests of the main results. **1)** Students in lower and upper secondary school are not fully comparable as grade repetition is more common among the latter. Excluding covariates (except age) leads to a reduction in the estimates for parents [OLS 0.91, se. 0.43]. This is consistent with socioeconomic factors correlating both with upper secondary grade repetition and the incidence of SARS-CoV-2. Dropping covariates leads to a small increase in the estimated impact for teachers [2.94, se 0.58] and their partners [1.58, se 0.71]. Both results are consistent with lower secondary partners being employed in more exposed occupations. Test have already shown poor balance when including parents of non-EU background. However, widening the sample to include these parents does not substantially alter the results. The OLS estimates with controls [1.09, se 0.42] and without controls [0.90, se 0.42] are similar to those for the main sample (SI Appendix, Table S3). **2)** Media searches reveal that some lower secondary schools closed spontaneously and preemptively, albeit for brief periods of time (see SI Appendix). As privately run independent schools were over-represented in this group, we exclude individuals connected to such lower secondary schools. This results in somewhat larger estimates for parents [1.33, se 0.46] (SI Appendix, Table S4), consistent with balancing tests reflecting high socioeconomic status and hence less predicted exposure among these parents (SI Appendix, Fig. S6a). Dropping independent lower secondary schools only slightly affects the estimates for teachers [2.63, se 0.63] and their partners [1.64, se 0.77] (SI Appendix, Table S5). **3)** It may have been more common among vocational programs to let small groups of students return to school to complete practical assignments. We therefore exclude parents connected to vocational upper secondary programs. These tend to be of lower socioeconomic status which is reflected in a poorly performing balancing test (SI Appendix, Fig. S6a). Consistent with this test, the point estimate is reduced [0.64, se 0.53] (SI Appendix, Table S4). **4)** Rather than controlling for employment in the healthcare sectors, we drop teacher households where the partner is a healthcare employee. As expected, the results remain unchanged (SI Appendix, Table S5). **5)** We derive a slightly different measure of parental exposure to lower secondary schools that allows for a broader sample of parents and the results are similar [0.98, se 0.34] (SI Appendix, Table S4). **6)** We broaden the comparison between lower and upper secondary parents by pooling those exposed to school years 8-11 and 7-12. This risks conflating the impact of exposure to open schools with student age. The estimate is lower for the 8-11 comparison [0.79, se 0.31] and even lower, and insignificant, for years 7-12 [0.20, se 0.26] (SI Appendix, Table S4). **7)** Household size might affect the risk of infection and it is decreasing by school year. Controlling for household size does however not affect the point estimates (SI Appendix, Table S6). **8)** To ensure that the results are not sensitive to the choice of cutoff dates, we use March 25 and April 16 for all groups. Since fewer cases are detected from the latter date, the OLS estimates are slightly reduced but the odds ratios are close to identical (SI Appendix, Table S7).

### Heterogeneity

How school closures affect the transmission of the virus depend on how they reduce contact between those potentially infected. This may differ depending on contextual factors and we analyze two types of heterogeneity. First, we allow the estimates for exposure to lower secondary schools to differ by population density in the district of residence. Second, since the timing of NPIs may affect their effectiveness (Caselli, Grigoli, Lian & Sandri, 2020), we let estimates vary by the regional rate of infections prior to school closure. The results in SI Appendix, Table S8 reveal interaction terms with large standard errors, not allowing a clear interpretation.

### Distribution of cases across schools

Past coronavirus outbreaks (SARS and MERS) have shown large individual variation in infectiousness implying that some individuals infected large number of secondary cases leading to ‘super-spreading events’ (Lloyd-Smith, James O and Schreiber, Sebastian J and Kopp, P Ekkehard and Getz, Wayne M, 2005). Estimates of the dispersion factor k – indicating heterogeneity in infectiousness – for SARS-CoV-2 vary, but suggest that this virus as well might spread in clusters (Endo, Abbott, Kucharski & Funk, 2020; Riou & Althaus, 2020). If the spread is highly clustered and the virus spread at the schools, we would expect most of the cases to be concentrated to a few schools.

The data at hand is not ideal to study such transmission patterns as the paucity of testing means that a large number of cases goes undetected. With this caveat in mind, Fig. S7 in SI Appendix shows how the cases are distributed across schools with different number of cases and Fig. S8 shows how cases are clustered in time within schools, separately for teachers and parents. There is some indication that cases among lower secondary school teachers were relatively concentrated, but among parents the cases are more evenly spread across schools and over time.

### Students

We do not study the impact of school closures on students, but for descriptive purposes Table S9 in SI Appendix shows estimates of infection rates for students under age 18 in school years 7–10. The incidence for students in year 10 is 0.53 PCR confirmed cases per 1000 and estimated differences between school years are not statistically significant. Because of age related differences in access to testing (see supplement), the severity of symptoms, risk behavior and patterns of socialization, results for students are likely to be biased and difficult to interpret. It can be mentioned that there were zero COVID-19 deaths recorded in age groups 2–19 in Sweden until late July, 2020. The rate of severe cases was also low; 94 hospitalizations were recorded among the 1.23 million students in compulsory school age (7–16) and 84 among the 339 000 youths in ages 17–19 (SI Appendix, Table S10). There might be other health implications for children and adolescents, but analyzing this is beyond the scope of this study.

## 3. Discussion

On March 18, 2020, upper secondary schools in Sweden moved to online instruction while lower secondary schools continued instruction as normal. This partial school closure provides a rare opportunity to study the impact on the transmission of SARS-CoV-2 during a period of widespread contagion. The impact of school closures on the transmission of the virus in society is best captured by the results for parents. We find that parental exposure to open rather than closed schools is associated with a somewhat higher rate of PCR-confirmed SARS-CoV-2 infections [OR 1.17; CI95 1.03–1.32]. The association is weaker for COVID-19 diagnoses from healthcare visits [OR 0.94; CI95 0.77–1.14] and severe cases that include hospitalizations and deaths [OR 0.84; CI95 0.64–1.11].

The positive association for PCR-confirmed cases could partly reflect other behavioral or biological differences between households with slightly younger and older children, but if treated as a causal the estimates indicate that a hypothetical closure of lower secondary schools in Sweden would have resulted in 266 fewer detected cases among the 253 538 parents in our sample. Limited testing capacity means that this only reflects a fraction of the actual number of cases, but it corresponds to a 15 percent reduction of the 1825 detected cases among lower secondary parents until mid-June (1072 cases when excluding healthcare workers). Since sample restrictions are made, the actual number of parents exposed to lower secondary schools is around 450 000 parents. The results thus indicate that closing lower secondary schools would have resulted in a 17 percent decrease in infections among 4.5 percent of the Swedish population. It is important to note that this captures both primary and secondary infections among household adults, and the full implications for virus transmission have to be derived using modelling. Although not conclusive in this regard, results are consistent with parental risk of infection increasing in student age. We might therefore somewhat underestimate the actual impact of keeping lower secondary schools open. More importantly, this means that the implications of keeping upper and lower secondary schools open may not be symmetric.

Teachers were more severely affected by the decision to keep lower secondary schools open. We estimate a PCR-confirmed infection rate twice as high among lower secondary teachers relative to teachers at upper secondary level [OR 2.01; CI95 1.52–2.67]. This is fully consistent with the results for COVID-19 diagnoses from healthcare visits [OR 2.01; CI95 1.45–2.79] and severe cases [OR 2.15; CI95 1.41–3.29]. When excluding healthcare workers, a comparison of SARS-CoV-2 infection rates across 124 occupations shows that upper secondary teachers are at the median while lower secondary teachers constitute the 7th most affected group. Other occupations with high infection rates (e.g. taxi drivers, driving instructors, social assistants, police officers) tend to have close interactions at work. This suggests that infections occur at school and there are some indications of clusters of cases among teachers. However, we cannot determine to what extent this is due to infections from students to teachers or if they reflect interactions between teachers. Primary school teachers had lower rates of infection than teachers at the lower secondary level and the patterns are consistent with teacher risk increasing in student age. Alternative explanations, such as different modes of interactions between the teaching staff, are possible, and this highlights that the impact of keeping schools open may not be symmetric across educational settings.

Increased infections among lower secondary teachers spill over to their partners who have a higher PCR-confirmed infection rate than their upper secondary counterparts [OR 1.29; CI95 1.00–1.67]. As for parents, the estimates are lower for COVID-19 diagnoses [OR 1.04; CI95 0.70–1.52] and severe cases [OR 1.09; CI95 0.62–1.92] among teachers’ partners.

Combining the estimates, 148 fewer cases of SARS-CoV-2 had been detected among lower secondary teachers (110) and their partners (38) if lower secondary schools had closed. To this we can add an estimate of 472 fewer cases among 450 000 adults exposed to lower secondary students in their households. Most transmission is within households so even if 620 fewer detected cases is a lower bound, this can be seen as relatively low compared to the country total of 53 482 detected cases until mid-June (35 556 excluding healthcare workers). Based on an age-specific case fatality rate (CFR) of 1.1% (SI Appendix, Table S10), this corresponds to 6.5 fewer deaths, 5 among parents and 1.5 among teachers and their partners. This counterfactual inference regarding mortality is highly uncertain, however. In our sample we count a total of 11 COVID-19 related deaths at the lower secondary level (9 parents, 1 teacher, 1 partner). The corresponding number at the upper secondary level is 16 (all parents). For severe health outcomes, we find 79 cases among 39 446 lower secondary teachers. According to the estimates, this number had been down to 46 if lower secondary schools had closed.

Closing the schools is a costly measure with potential long-run detrimental effects for students. The results presented are in line with theoretical work indicating that school closure is not an effective way to contain SARS-CoV-2 (Viner et al., 2020), at least not when facing as high a level of contagion as Sweden did during the spring of 2020. It is not clear how the results generalize to other settings and studies have found both positive and negative associations between closed schools and the rate of transmission of SARS-CoV-2 (Hsiang et al., 2020; Auger et al., 2020; Haug et al., 2020; Isphording, Lipfert & Pestel, 2020). The mixed evidence could reflect methodological differences and difficulties isolating the impact of schools. However, they could also reflect differences in how schools are organized and local conditions at the time of intervention. Unfortunately, our results do not allow any firm conclusions regarding interactions between school closures and local conditions. Another potentially important difference between settings is the level the precautionary measures undertaken within schools. According to an international comparison (Guthrie et al., 2020), the measures recommended in Sweden (Public Health Agency of Sweden, 2020b) are best described as mild. In particular, there is no quarantine of those exposed unless they show symptoms of infection, no imposed class size reductions, and face masks are rarely used (YouGov, 2020).

While the overall impact on overall virus transmission was limited according to this study, keeping lower secondary schools open had a quite substantial impact on teachers and the results suggest that the risk to teachers can be increasing in student age. This should be taken into account and precautionary measures could be considered.

## 4. Materials and methods

We construct estimation samples for parents, teachers and their partners using registers held by Statistics Sweden. Through the Multi-Generation register (MGR) per December 31 2019 and Longitudinal integrated database for health insurance and labor market studies (LISA) per December 31 2018 we identify all parents with children in relevant ages in their households. Children are assigned to school year, schools and upper secondary programs using the Student Register as per October 15, 2019. We sample all parents in Sweden and their partners living in households with the youngest child in lower or upper secondary school. We also include parents with a biological or adopted child who do not live in the same household but in the same region. The main analysis excludes parents born outside Sweden, the Nordic countries and the EU. Information on detailed place of residence as of December 31, 2019, is available for all individuals in Sweden in the Register of the Total Population (RTB). The sample of teachers includes all teachers working at the lower or upper secondary levels in the Teacher Register and refers to the status of the teacher in the fall of 2019. Their partners are identified using the household identifier in LISA. See the supporting information for further details on the estimation samples. Information on the covariates: disposable income, educational attainment and occupation are available in LISA. Occupations are reported according to the Swedish Standard Classification of Occupations (SSYK 2012) which is based on the international classification (ISCO-08). There are 46 occupation categories on the 2-digit level.

Information on positive PCR tests of SARS-CoV-2 is from the Swedish Public Health Agency. Up until late July there were 75 933 reported cases of SARS-CoV-2, out of which test dates are missing for 2506 cases. As majority of the cases without test date are reported outside the main period of analysis, they are discarded. Personal identifiers are available for all cases, making it possible to link the test results to register data. Information on COVID-19 diagnoses until June 30 from the Inpatient-and Outpatient register is available from the National Board of Health and Welfare and on deaths from the Cause of Death register held by Statistic Sweden. By June 30, 2020, 33 596 individuals had been diagnosed with COVID-19 (ICD 10 codes U07.1 or U07.2) either in the Patient registers or the Cause of Death register.

Table 2 reports descriptive statistics for parents, teachers and teachers’ partners, starting with the incidence of positive PCR tests of SARS-CoV-2 as of June 15. Since healthcare workers were prioritized for testing, we also present the incidence excluding those working in healthcare. Healthcare workers are excluded by dropping those with occupational codes 15, 22, 32 and 53 (SSYK2012). The table further shows the incidence of positive PCR test prior to the cut-off date chosen to reflect the infection rate prior to the move to online instruction at the upper secondary level (March 25 for teachers, April 1 for parents and partners). The table next displays the incidence of COVID-19 diagnoses from healthcare visits and the incidence severe cases as of June 30. Finally it displays the number of COVID-19 related deaths in each sample as of July 25 and the number of individuals in each group.

We use ordinary least squares (OLS) and logistic regressions (Logit) to empirically analyze if SARS-CoV-2 infection can be attributed to being exposed to open or closed schools. We estimate the following OLS regression model for the three populations: parents, teachers and teachers’ partners:

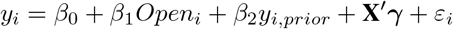

The outcome *y*_*i*_ is an indicator variable for a positive SARS-CoV-2 PCR test or being diagnosed with COVID-19 by a doctor in outpatient care or at a hospital. There is just one positive test per individual and *y*_*i*_, *prior* is an indicator for SARS-Cov-2/COVID-19 before the cut-off date. Including *y*_*i*_, *prior* is a way of excluding pre-period cases without dropping such observations. *Open* is an indicator variable taking the value (1) if individual *i* is exposed to (open) lower secondary schools. Parents with the youngest child in lower secondary school are defined as exposed and parents with the youngest child in upper secondary school are defined as unexposed. Lower secondary teachers and their partners are defined as exposed and their upper secondary counterparts as unexposed. **X** is vector of individual and household characteristics. When estimating the model for teachers the vector includes: 20 indicators for age categories, sex, 7 indicators for categories of educational attainment, 46 indicators of categories of partners’ occupation, 12 region indicators of country of origin for those not born in Sweden, log of household income, indicator of having a teaching position, percent of full time position, 290 indicators of municipality of residence and household exposure to the number of children in age groups 2–6, 7–16, 17–19, and 20+ who reside in the same region as the teacher. The municipality fixed effects are exchanged for 21 region fixed effects when estimating the logistic model. The equivalent vector of variables is used for teachers’ partners, with the exception of own occupation instead of partner occupation. The vector of controls for parents include a similar set of variables as for teachers: age categories, sex, municipality of residence, educational attainment, occupational categories (own and partners’), region of origin for those not born in Sweden (3 indicators in the main sample), the log of disposable family income and indicators for missing data on any of these variables. Migrant from non-EU/Nordic countries are excluded from the main sample of parents. Standard errors are clustered at the school level when estimating the model for teachers and teachers’ partners and at the household level when studying parents.

## Data Availability

The data used for the analysis have been obtained by combining several administrative registers collected and held by three government agencies: Statistics Sweden, The National Board of Health and Welfare and The Public Health Authority. Access to these data sets is restricted by national regulations, but any researcher wishing to replicate our results can apply for access through these agencies. Since the study involves sensitive individual level data, getting access to the data also requires approval from an Ethics Board.

## Supplementary information

### The development of the pandemic and non-pharmaceutical interventions

The first case of SARS-CoV-2 in Sweden was reported on January 31, 2020, and the disease was classified as a danger to public health and to society on the following day (Public Health Agency of Sweden, 2020a). Among other things, this classification means that all documented cases of active infection have to be reported to the Public Health Agency. The first death from COVID-19 occurred on March 11. The daily number of deaths increased rapidly and peaked in the first half of April whereafter the daily number of deaths declined gradually. By the end of the school year in mid-June, the 7-day average of daily deaths was around 30 and the cumulative number of deaths 5140 (51 per 100 000 inhabitants).

The hardest hit region in both absolute and relative terms was the Stockholm region with 2.4 million inhabitants. Stockholm recorded 2211 deaths (93 per 100 000) and 16 275 cases (685 per 100 000) by mid-June. In deaths per 100 000 inhabitants, Stockholm was followed by Sörmland (79), Västmanland (55) and Dalarna (52). The second largest region of Sweden, Västra Götaland, had by June 15 reported 649 deaths among its 1.7 million inhabitants. Testing scaled up faster in this region than in Stockholm and the total number of cases was 11 000. The region of Skåne with 1.4 million inhabitants was less affected and reported 16 deaths per 100 000 and a total of 2300 cases by mid-June.

The Swedish Public Health Agency introduced several measures to reduce the transmission of the virus (Public Health Agency of Sweden, 2020c). On March 10, a recommendation against unnecessary visits to care facilities was issued and on March 11 public gatherings of more than 500 people were banned. On March 13, people were recommended to stay at home when having symptoms of illness and those who could work from home were recommended to do so on March 17. On March 18, upper secondary schools and institutions of higher education moved to online instruction. On March 19, a recommendation against unnecessary travel was issued and on March 24, restaurants and bars were instructed to increase the distance between costumers. Public gatherings above 50 persons were banned on March 27 and visits to elderly care facilities were banned the following day. On April 1, stricter recommendations on social distancing for the public were issued. On June 13, the recommendation against unnecessary travel was lifted. Throughout the period, there was no official recommendation that those without symptoms should stay at home, even if the household was shared with individuals with confirmed SARS-CoV-2 infection.

Mobility both within and between Swedish regions declined substantially as a response to the pandemic and the recommendations issued by the authorities (Public Health Agency of Sweden, 2020d). The distance individuals moved from their homes during a day was substantially reduced and the decline in mobility was similar for residents in areas with different socioeconomic and demographic characteristics (visible minorities, highly educated, poor, and being 70 years or older) (Dahlberg et al., 2020).

### Swedish schools during the pandemic

Compulsory schools (age 7–16) were kept open for instruction and to reduce transmission the following precautionary measures were recommended (Public Health Agency of Sweden, 2020b): enhanced facilities for hand washing and disinfection; posters encouraging hand washing; increased distance in classrooms and dining halls, if possible; avoidance of large gatherings, as far as possible; minimize activities like open houses and parental meetings; increased outdoor activities, if possible; avoidance of close contacts between staff and students and between students; enhanced cleaning of heavily exposed areas and keyboards/tablets. Compared to school opening policies in other countries, the precautionary measures in Sweden are best described as mild (Guthrie et al., 2020). In particular, there is no mandated quarantine of those exposed who do not show symptoms, no imposed reductions of class size and no recommendations concerning the use of face masks.

On March 18, upper secondary schools and institutions of higher education moved to online instruction. Upper secondary schools thus closed for normal instructions just as the number of deaths and ICU admissions began to increase (see Fig. 1 in the main text). Although upper secondary school moved to online teaching, some teachers were still teaching online from the school premises. According to a survey conducted by a large teachers’ union during the last week of April and first week of May, 21 percent taught from the school, 46 percent partly from home, and 33 percent only from home (National Union of Teachers, 2020). As expected, compulsory school teachers mainly taught from school; 2 percent of the teachers in compulsory schools had been partly teaching online from home and 1 percent had only been teaching from home. There have also been media reports of substantial student absenteeism in compulsory schools. Again there are no official reports but according to the same survey, 18 percent of compulsory students were absent on a typical day. In a survey of 27 compulsory schools conducted by the National Board of Education during late April, 7 schools reported that absenteeism among compulsory school students was about normal, 13 that there was an increase in absenteeism of between 20 and 50 percent, and 7 stated an increase of more than 50 percent (Swedish National Agency for Education, 2020c). The conclusion drawn from this survey is that student absenteeism increased, but not dramatically so.

### Data and sample restrictions

The sample of parents is constructed as follows. We define household adults who are exposed to their own children (biological or adopted) or a new partner’s children from a previous relationship as parents. For separated parents, we use the household identifier in LISA to identify any current partner. This enables us to identify new couples who are either married or have common children. Households consisting of unmarried cohabitant couples without common children cannot be identified and will be categorized as single households. The study population consists of parents who have children in school years 7–12 in the household, or biological children in these school years living in the same region. Because parents are less likely to interact regularly with children living at a distance, only children residing in the same region are considered in the analysis. There are 21 regions in Sweden and they are thus relatively large geographical areas. There were also recommendations against leaving the region of residence during most of the spring 2020.

We sort parents by the age of the youngest child connected to the parents in the household or through biological links. Parents are considered exposed to lower secondary schools if their youngest child is enrolled in school years 7–9. Unexposed parents are defined by their youngest child being enrolled in upper secondary school. In the analysis we focus on parents with the youngest child in school years 9 and 10 since they are likely to be the most similar in other aspects, except for parents with their youngest child in the 9th school year being exposed to an open school. We further exclude those born outside of Sweden, the Nordics, and the EU. After this restriction, the main sample consists of 166 630 parents connected to school years 9 and 10. 480 291 parents are connected to school years 7 though 12.

The teacher sample consists of teachers working in lower and upper secondary schools according to the Teacher Register. Teachers with children born in 2019 are excluded as they are likely to be on parental leave during the spring of 2020. We also exclude those recorded as being on leave of absence during the fall of 2019. The final sample consists of 72 946 lower and upper secondary teachers. In a descriptive analysis we include lower and upper primary school teachers (school years 1–6) identified in the Teacher Register. When including these, the teacher sample consists of 137 213 individuals. For the sample of partners to lower and upper secondary teachers, we connect partners to teachers using the household identifier from LISA. This enables us to identify partners who are either married to or have common children with the teacher. The resulting sample consists of 47 383 partners.

Our main outcome variable is positive PCR tests reported to the Public Health Agency but we also analyze the incidence of COVID-19 diagnoses from healthcare visits and severe cases of COVID-19 (hospitalizations and deaths) reported to the National Board of Health and Welfare. The first case of SARS-CoV-2 in Sweden was reported on January 31, 2020, and the disease was classified as a danger to public health and to society on the following day (Public Health Agency of Sweden, 2020a). Among other things, this classification means that all documented cases of active infection have to be reported to the Public Health Agency. Testing capacity was slow to expand and from March 13 (week 11), testing was directed towards healthcare employees and individuals with symptoms of COVID-19 in need of healthcare. As shown in Fig. S1, testing increased substantially from early June (week 23). Healthcare is the responsibility of Sweden’s 21 healthcare regions as is testing for SARS-CoV-2. Thus, there are regional differences in testing capacity as well as rules and recommendation regarding testing. Some regions have recommended not to test children under 16 (for example Västra Götaland and Uppsala) and some have not had any age restrictions (for example Skåne). The number of detected cases does therefore not well reflect the actual rate of infections and the rate of positive tests remained high throughout June (week 27). By June 15, a total of 383 000 PCR tests had been performed (3 800 per 100 000 inhabitants) (Public Health Agency of Sweden, 2020e).

### Covariate balance

For estimation of the causal effect on parents the identification strategy hinges on the similarity of parents with their youngest child in school years 9 and 10. Apart from a 1-year age difference, these groups should be balanced on covariates in order to be valid counterfactuals. We test this assumption by showing balancing tests where we predict the SARS-CoV-2 test outcome using the observable covariates (apart from parental age) of parents with the youngest child in school years 7–12. Fig. S2 shows covariate balance for the main sample of parents. The corresponding balancing test when non-EU migrants are included is shown in Fig. S3. Odds ratios for both samples of parents without controlling for covariates are shown in Fig. S4 and odds ratios with covariates are shown in Fig. S5. The OLS estimates for both samples of parents, with and without covariates, are shown in Table S3. Table S3 also shows estimates for teachers and teachers’ partners, with and without covariates.

### Results including primary school teachers

We extend the population of teachers at open schools to include lower (school years 1–3) and upper (school years 4–6) primary school teachers. Results for confirmed PCR-tests and COVID-19 diagnoses when controlling for covariates are shown in Table S2.

### Additional results and robustness tests

The propensity to get tested for SARS-CoV-2 could be affected by being connected to open and closed schools, regardless of health status. This is less of a concern for COVID-19 diagnoses made by the healthcare sector, especially severe cases which require hospital care or cause death. Results for severe cases, defined as admittance to hospital or death due to COVID-19, are presented for all groups in Table S1.

Some lower secondary schools spontaneously moved to online instruction and may thus be classified as having on-site instruction when they in fact conducted the teaching online. No official records on such closures exist but media searches reveal that they were rare and short-lived (see below). Privately managed independent lower secondary schools are over-represented in reports on proactive closures and we therefore exclude such schools as a robustness test. Students attending independent schools are generally from a more advantaged socioeconomic background and excluding them introduces imbalance to the sample of parents (Fig. S6). OLS estimates excluding independent lower secondary schools for parents are shown in Table S4. Corresponding results for teachers and their partners are shown in Table S5.

Upper secondary schools were allowed to let small groups of students complete practical elements of education and assignments, provided that this could be done safely (Swedish National Agency for Education, 2020b). Such practices may have been more common at vocational programs and as a robustness test we exclude parents exposed to such upper secondary programs. This amounts to excluding parents of relatively disadvantaged socioeconomic background, which means that the exclusion introduces imbalance among parents (Fig. S6). OLS estimates imposing this exclusion are shown in Table S4.

The baseline specifications controls for the occupation of teachers’ partners. As a robustness test, we instead drop the teachers and partners who are exposed to the healthcare sector through the partners’ occupation (occupational codes 15, 22, 32 and 53). The results are shown in Table S5.

We use an alternative measure of exposure to lower secondary school for parents. Parents are then defined as exposed if they have a child in the household, or a child residing in the same region, in lower secondary school. Families with children too old to be in secondary school are dropped, as are families whose youngest child attends school below year 7. We control for having a child in school years 11 and 12 and the results presented in Table S4 thus shows the impact of being exposed to a child in lower secondary school compared to being exposed to a child in upper secondary school year 10. Table S4 also shows results where we pool parents with the youngest child in school years 8–11 and 7–12.

Household size tends to decrease in student age and Table S6 shows results for parents when controlling for this variable. Table S7 presents the sensitivity to using the cut-off dates March 25 and April 16 for parents, teachers and teachers’ partners.

### Heterogeneity analysis

The expected impact of school closures on virus transmission depends mainly on the magnitude of contact reduction. Two factors that may be of importance for the effect is population density and how widely spread the virus was prior to schools closing. A study of US districts show that transmission of SARS-CoV-2 increases with population density (Korevaar et al., 2020). To investigate this matter we implement a heterogeneity analysis by district population density, categorizing districts with a population density above the 75th percentile as high density districts.

Timing has been shown to be important for the effectiveness of NPIs (S. Lai et al., 2020). We therefore investigate whether the impact of school closures depends on the level of virus transmission prior to school closure. Regions with above the populated weighted median spread of 12 cases per 100 000 are categorized as high spread regions, i.e. the regions (cases per 100 000 in parenthesis): Stockholm (20), Uppsala (16), Ö stergötland (16), Skåne (16), Sörmland (13) and Jönköping (12).

The econometric model is modified by adding interaction terms between indicators for high population density, respective high initial contagion, and exposure to lower secondary school as well as interactions with all control variables except for the municipality indicators. The results are reported in Table S8.

### Distribution of cases across schools

Although limited by the low testing rate, an illustration of the aggregation of cases across schools and over time can provide some evidence of the role of super-spreading events. To investigate whether there is substantial heterogeneity across schools, we aggregate cases across schools for parents and teachers, respectively. Cases among parents connected to a school through students in school years 7–12 are aggregated to the school level which means that cases among parents to several children are connected to more than one school. When excluding schools with less than 50 connected parents there are 1455 lower and 1149 upper secondary schools in the data. Among these schools, 25 percent of upper secondary and 32 percent of lower secondary schools had no cases. Since upper secondary schools on average are larger (397 connections compared to 312 for lower secondary schools) we mechanically expect more cases in upper secondary schools. Fig. S7 shows the fraction of total cases in lower respective upper secondary schools with one to 28 cases. For both types of schools, a majority of cases occurred at schools with few cases. To analyze how the cases are clustered over time, we aggregate the cases into episodes. If all cases within a school occur the same or adjacent week it is coded as one episode. If cases are more dispersed over time, the school is coded as having more than one episode of infection outbreaks. Fig. S8 displays the fraction of schools with at least two cases in total that have one or more than one episodes. The pattern is similar for lower and secondary schools, with about 60 percent of the schools having one outbreak episode and 40 percent having more than one episode.

We conduct the same analysis for teachers at schools with more than 5 teachers. As for the analysis of parents, there are no cases in a majority of schools (90 percent of lower secondary and 93 percent of upper secondary schools). Moreover, most cases are recorded in schools with only one case (Fig. S7). Among upper secondary schools there are no schools with more than two cases, whereas among lower secondary schools there are some schools with three or more cases. The main analysis shows that keeping lower secondary schools open resulted in approximately 100 additional cases among lower secondary school teachers. According the patterns of distribution presented here, about a third of these can be found in schools with many cases and two thirds in schools with only one case. Turning to the analysis of outbreak episodes there is some indication of more clustering of outbreaks among teachers in lower secondary than upper secondary schools (Fig. S8).

### Students

We show descriptive results of infection rates for students by school year in Table S9. Due to the discussed age restrictions for testing and risk of differing behavior for students over 18, we show results for students below age 18 in school years 7–10. As with parents and teachers, we control for observable characteristics such as gender, region of origin, and mother and father log disposable income, occupation, region of origin, education, missing values, and number of siblings in different age groups. We restrict attention to students with parents born within the EU and Nordics due to balancing of covariates concerns.

### Media searches

In order to get information on spontaneous closures of lower secondary schools, media searches were conducted using the service Mediearkivet/Retriver and on Sveriges Radio’s web page (public service radio with substantial local presence). Search terms were permutations of “school closure” (skolstängning/skola stängd), “distance education” (distansundervisning), “online education” (onlineundervisning), “corona” and “covid”. Results for individual schools were followed by web searches to find more information on each particular case. Spontaneous closures were recorded as proactive if they did not occur as a result of cases detected at the school and reactive otherwise. Provided that information is available, a closure is labelled as brief if the duration was less than a week.

In total, reports on 40 closures were found (27 among privately managed independent schools). 29 of these were proactive (22 among independent schools) while 11 were reactive (5 among independent schools). Spontaneous closures thus appear to have been rare and independent schools are vastly over-represented among those that closed proactively. Two of the reactive closures were on advice from the local disease protection officer and they both occurred late in the school year (June 6 and 8). Information on the duration was usually not available, but of the 18 reports from which the duration can be judged, 12 were brief. Several of the closures were also partial, meaning that school days were cut short, rolling schedules introduced, or that instruction partially moved online. Details on each specific report are available from the authors.

### Cases, deaths and the case fatality rate

To extrapolate the expected effect of school closure on the number of deaths in Sweden we derive the case fatality ratio (CFR) for different age groups. CFR is calculated by dividing the number of deaths with the number of cases and hence crucially depend on the testing regime. Table S10 shows the incidence of detected SARS-CoV-2 in different age groups until June 15, 2020, and the number of deaths among these cases reported until July 25. The numbers are shown both including and excluding healthcare workers, for which testing was more accessible. The CFR increases with age, except for the higher value for the youngest age group due to one dead child. This child was younger than one years old and thus not directly exposed to schools. The average age among teachers is 48, their partners 49 and parents 50 years old. Based on the CFR distribution in Table S10 we calculate the expected effect on mortality among lower secondary parents using a CFR of 1.1 percent.

**Fig. S1:**
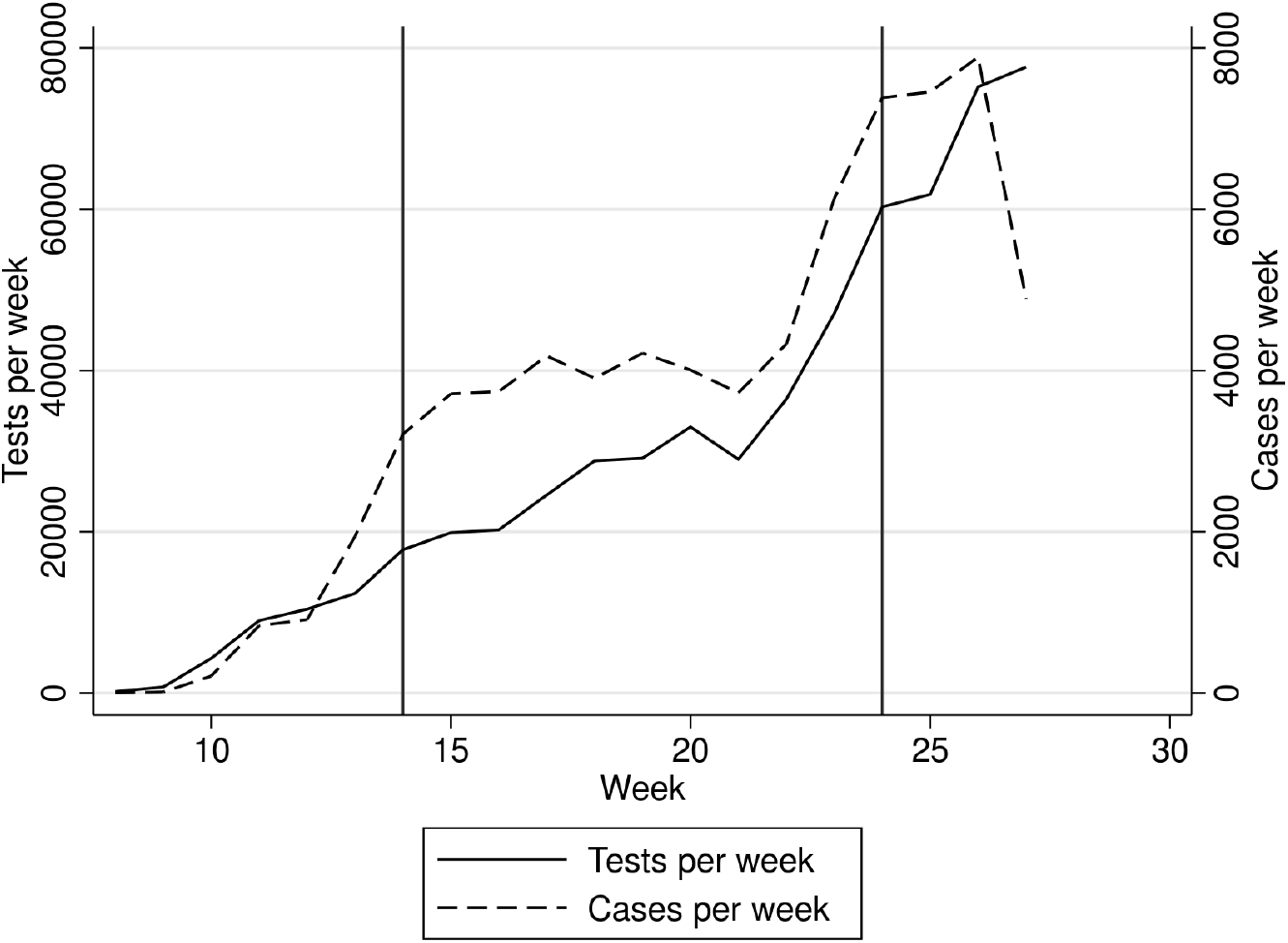
Tests and cases per week. Weekly number of PCR tests and positive cases. Vertical lines indicate weeks 14 and 24, the approximate period of analysis. Data from the Public Health Agency (Public Health Agency of Sweden, 2020e).

**Fig. S2:**
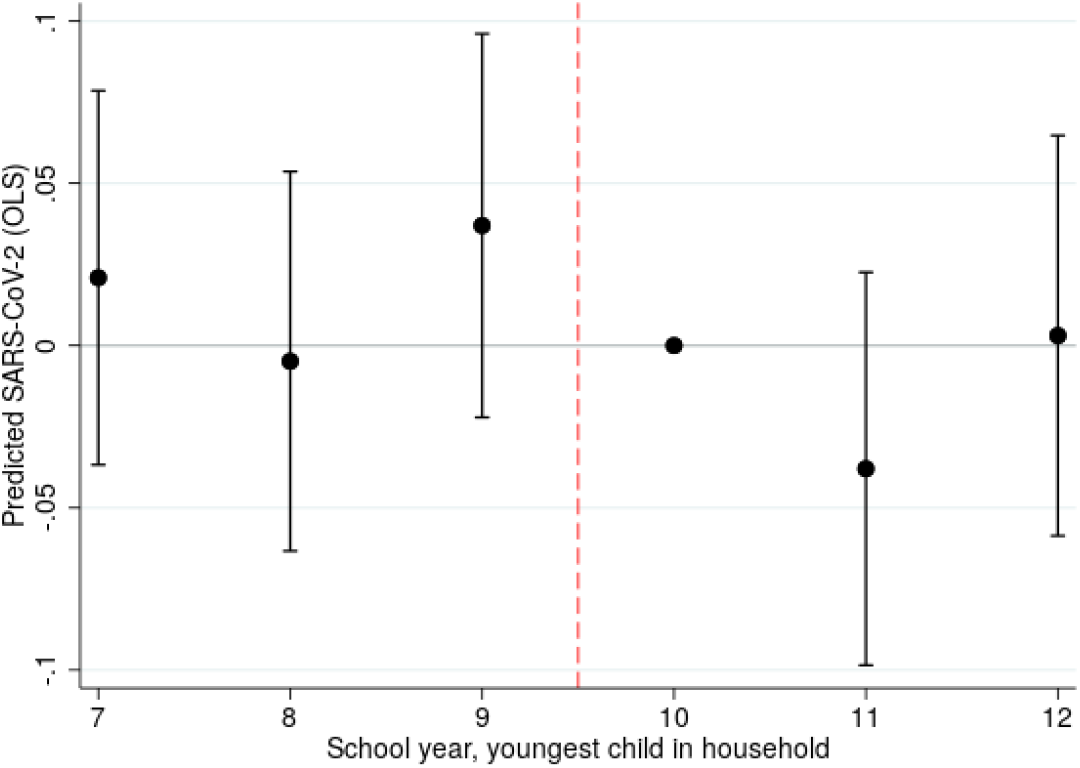
Covariate balance, main sample. Predicted SARS-CoV-2 regressed on school year of the youngest child in the household for parents born within EU and Nordics. Predicted outcome using sex, occupation, educational attainment, income, regions of residence and of origin for parents. The reference category is school year 10 and 95% confidence intervals are indicated.

**Fig. S3:**
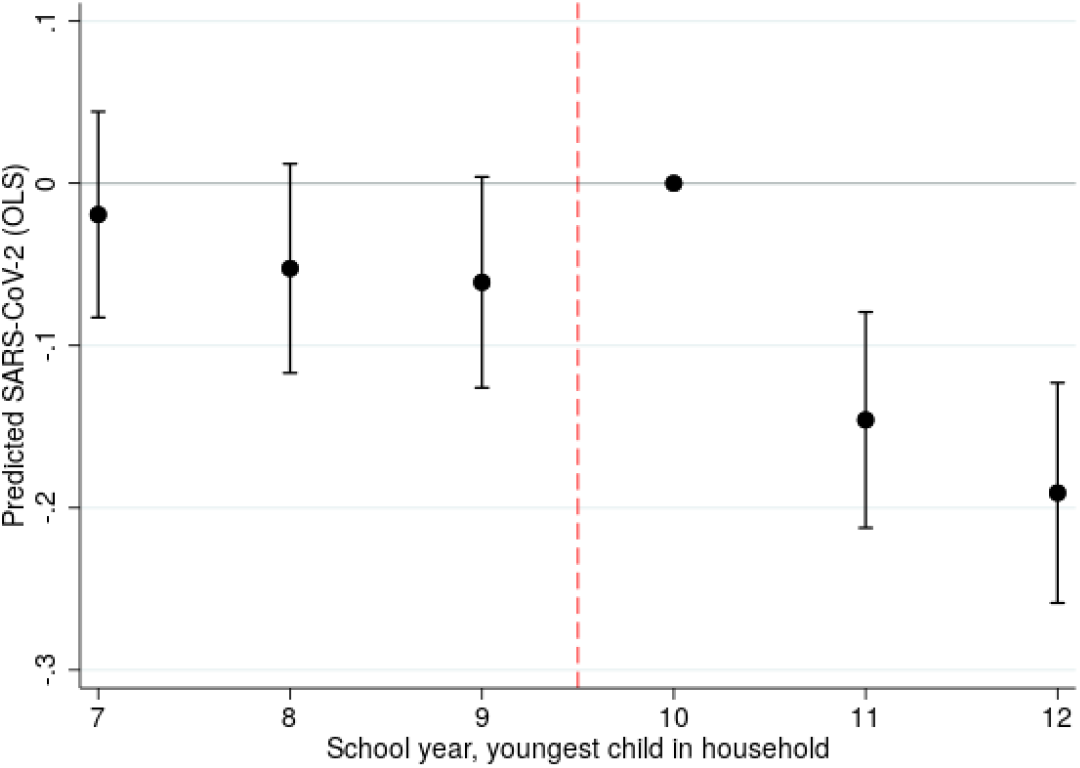
Covariate balance, all parents. Predicted SARS-CoV-2 regressed on school year of the youngest child in the household for all parents. Predicted outcome using sex, occupation, educational attainment, income, regions of residence and of origin for parents. The reference category is school year 10 and 95% confidence intervals are indicated.

**Fig. S4:**
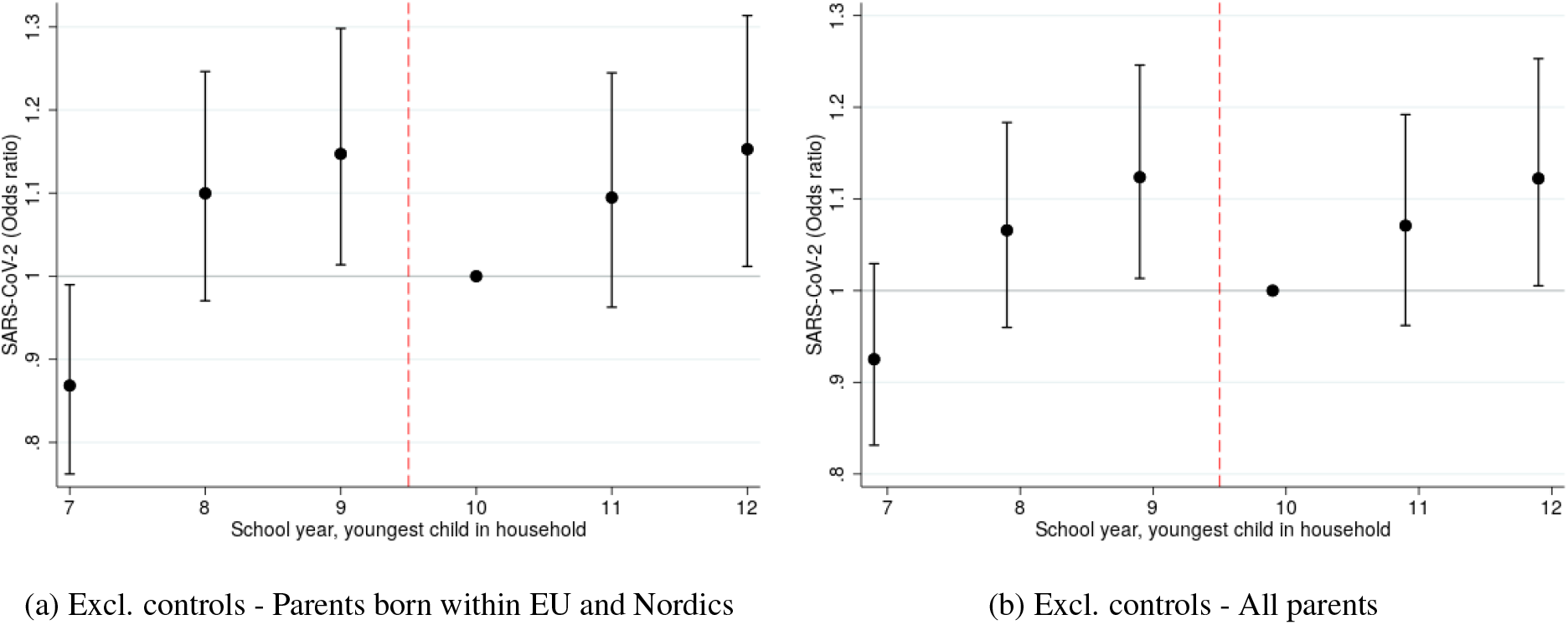
Results excluding covariates. SARS-CoV-2 odds ratios for parents by school year of the youngest child in the household excluding all control variables (except for age effects and *y*_*prior*_). Odds ratios estimated using logistic regression. The reference category is school year 10 and 95% confidence intervals are indicated. Fig. S4a shows outcomes including parents born within the EU and Nordics, which is our main study population. Fig. S4b shows outcomes including all parents.

**Fig. S5:**
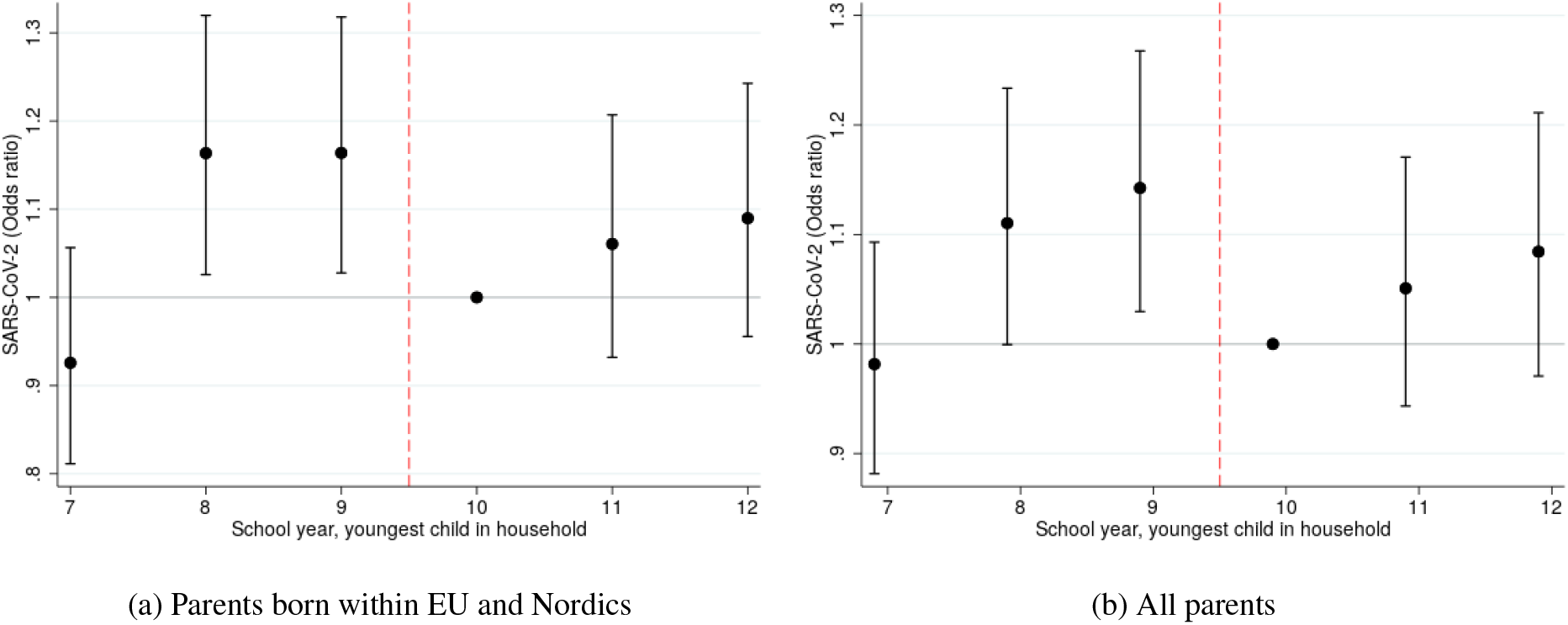
Results including covariates. SARS-CoV-2 odds ratios for parents by school year of the youngest child in the household. Odds ratios estimated using logistic regression. The reference category is school year 10 and 95% confidence intervals are indicated. Fig. S5a shows outcomes including parents born within the EU and Nordics, which is our main study population. Fig. S5b shows outcomes including all parents.

**Fig. S6:**
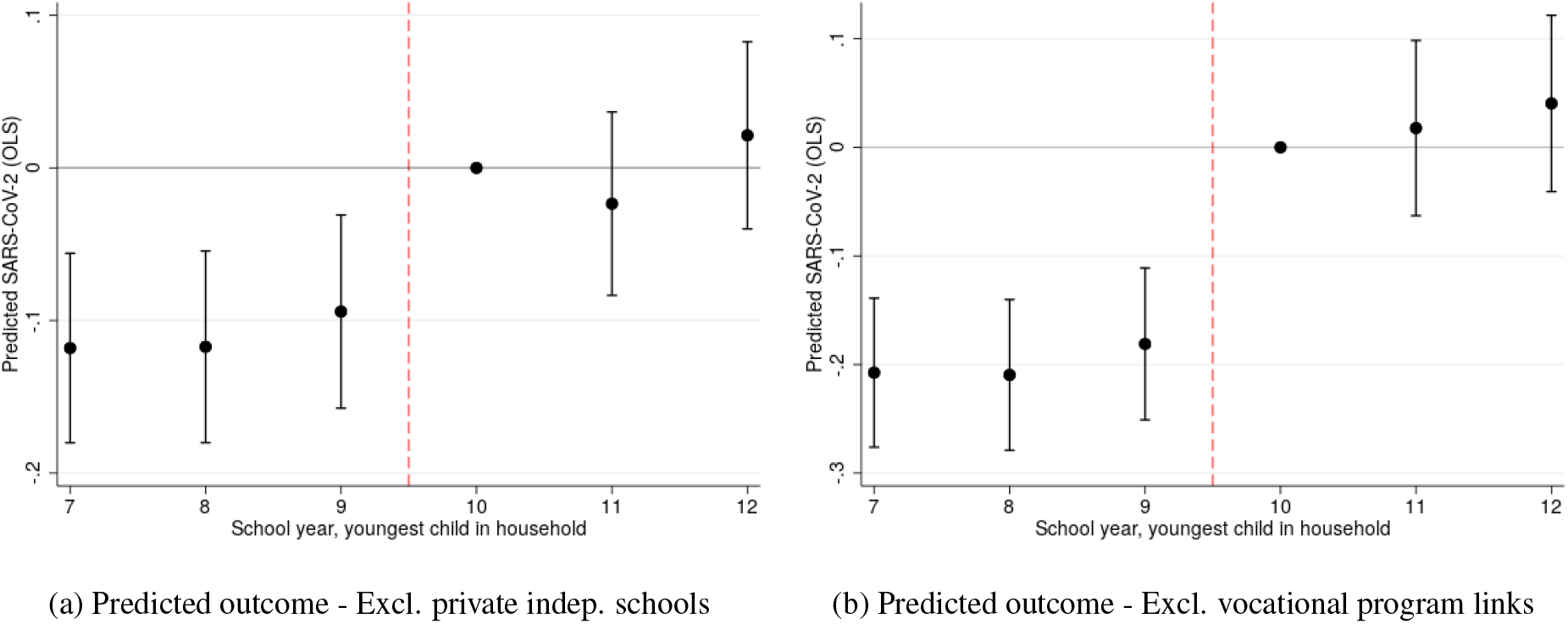
Covariate balance for subsamples. Predicted SARS-CoV-2 regressed on school year of the youngest child in the household for parents born within EU and Nordics, excluding private independent schools and vocational program links separately. Predicted outcome using sex, occupation, educational attainment, income, regions of residence and of origin for parents. The reference category is school year 10 and 95% confidence intervals are indicated. Fig. S6a shows outcomes excluding private independent school links. Fig. S6b excludes vocational program links.

**Fig. S7:**
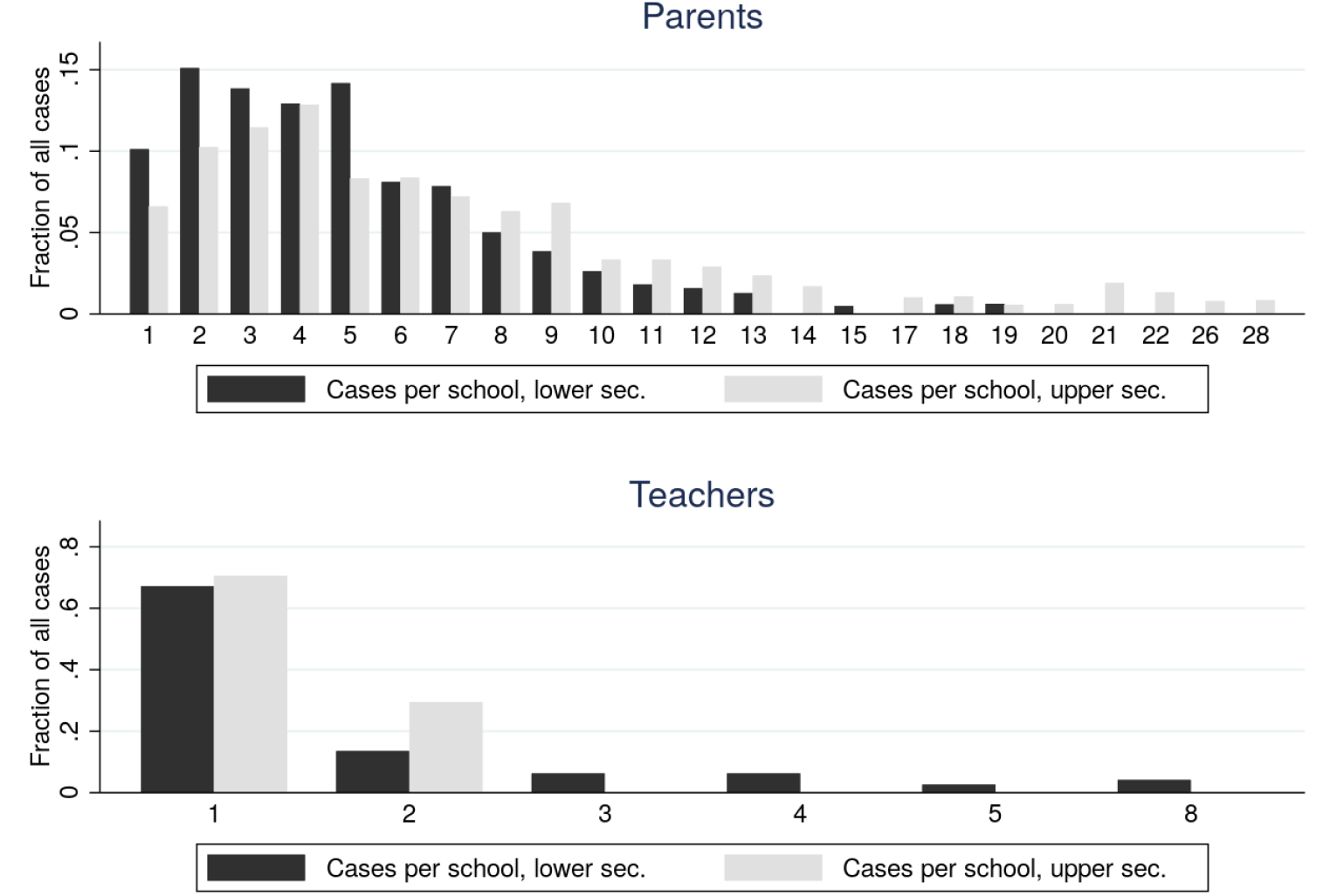
Distribution of cases across schools. The figure shows the share of total cases at schools with 1 to 28 cases.

**Fig. S8:**
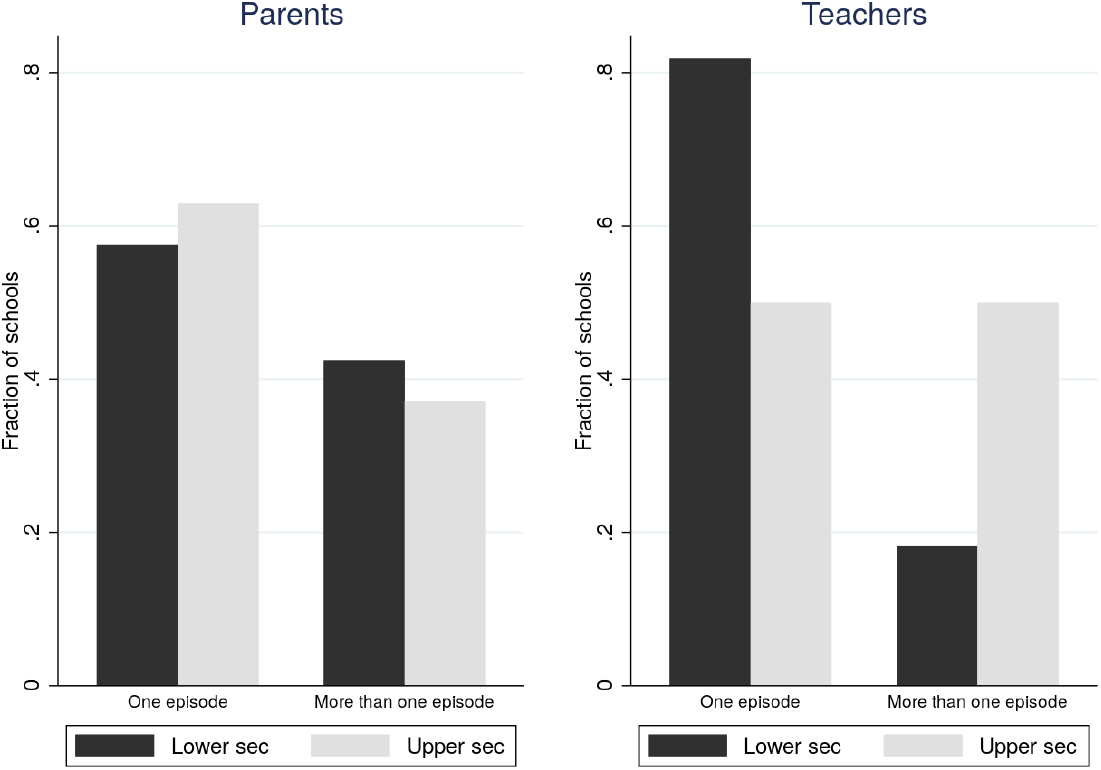
Episodes of cases within schools. The figure shows the share of schools with at least two cases which had all cases in one week or adjacent weeks and the fraction of schools with cases at least one week apart.

**Table S1:** Impact of exposure to open schools on PCR tests and severe COVID-19 diagnoses

|  | Parents |  | Teachers |  | Teachers' partners |  |
| --- | --- | --- | --- | --- | --- | --- |
|  | OLS (cases/1000) |  |  |  |  |  |
|  | PCR | Severe cases | PCR | Severe cases | PCR | Severe cases |
| Open school | 1.05**<br>(0.43) | -0.21<br>(0.18) | 2.81***<br>(0.59) | 0.84***<br>(0.28) | 1.47**<br>(0.71) | 0.08<br>(0.31) |
| Mean dep. var. | 6.37 | 1.40 | 2.96 | 0.96 | 5.10 | 1.01 |
| Obs. | 166,630 | 166,719 | 72,946 | 72,976 | 47,383 | 47,413 |
|  | Logit (odds ratios) |  |  |  |  |  |
|  | PCR | Severe cases | PCR | Severe cases | PCR | Severe cases |
| Open school | 1.17**<br>[1.03,1.32] | 0.84<br>[0.64,1.11] | 2.01***<br>[1.52,2.67] | 2.15***<br>[1.41,3.29] | 1.29*<br>[1.00,1.67] | 1.09<br>[0.62,1.92] |
| Obs. | 163,195 | 150,571 | 70,151 | 62,249 | 44,025 | 34,563 |

Note: \*\*\* $p < 0.01$ , \*\* $p < 0.05$ , \* $p < 0.1$ . Standard errors in parenthesis clustered the at the household level for parents and school level for teachers and partners. Severe cases include COVID-diagnoses registered at hospital or as death. The effects are estimated using linear probability models (OLS) and logistic regressions (Logit).

**Table S2:**
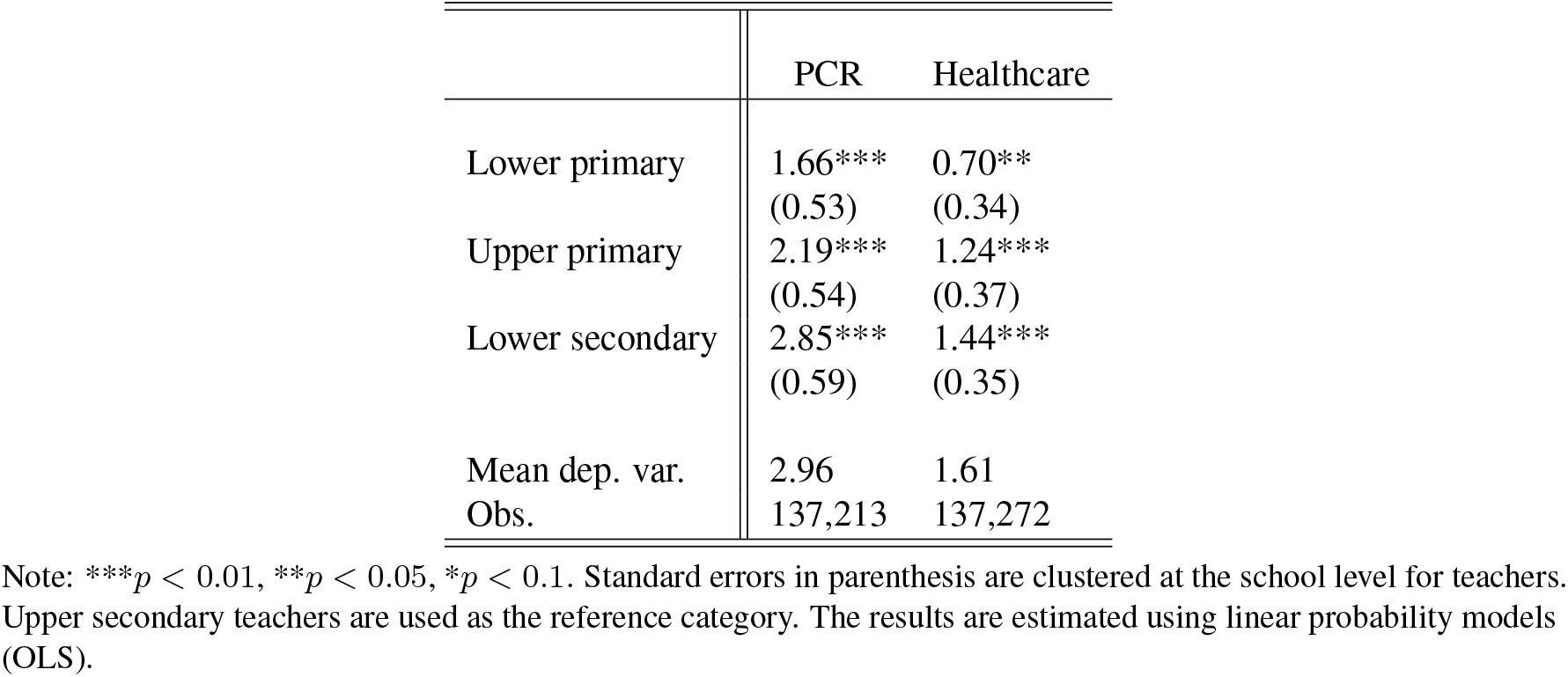
SARS-CoV-2 among lower primary, upper primary, and lower secondary teachers relative to upper secondary teachers (OLS)

|  | PCR | Healthcare |
| --- | --- | --- |
| Lower primary | 1.66***<br>(0.53) | 0.70**<br>(0.34) |
| Upper primary | 2.19***<br>(0.54) | 1.24***<br>(0.37) |
| Lower secondary | 2.85***<br>(0.59) | 1.44***<br>(0.35) |
| Mean dep. var. | 2.96 | 1.61 |
| Obs. | 137,213 | 137,272 |

Note: \*\*\* $p < 0.01$ , \*\* $p < 0.05$ , \* $p < 0.1$ . Standard errors in parenthesis are clustered at the school level for teachers. Upper secondary teachers are used as the reference category. The results are estimated using linear probability models (OLS).

**Table S3:** Main results for parents, teachers & partners - including and excluding controls. Outcome: Positive PCR tests per 1000

|  | Parents (main) |  | Parents (all) |  | Teachers |  | Partners |  |
| --- | --- | --- | --- | --- | --- | --- | --- | --- |
|  | Controls | Excl. controls | Controls | Excl. c. | Controls | Excl. c. | Controls | Excl. c. |
| Open school | 1.05**<br>(0.43) | 0.91**<br>(0.43) | 1.09***<br>(0.42) | 0.90**<br>(0.42) | 2.81***<br>(0.59) | 2.94***<br>(0.58) | 1.47**<br>(0.71) | 1.58**<br>(0.71) |
| Mean dep. var. | 6.37 | 6.37 | 7.58 | 7.58 | 2.96 | 2.96 | 5.10 | 5.10 |
| Obs. | 166,630 | 166,630 | 205,843 | 205,843 | 72,946 | 72,946 | 47,383 | 47,383 |

Note: \*\*\* $p < 0.01$ , \*\* $p < 0.05$ , \* $p < 0.1$ . Standard errors in parenthesis are clustered at the household level for parents and school level for teachers and partners. “Open school” is defined as exposure to lower secondary school. The results are estimated using linear probability models (OLS).

**Table S4:**
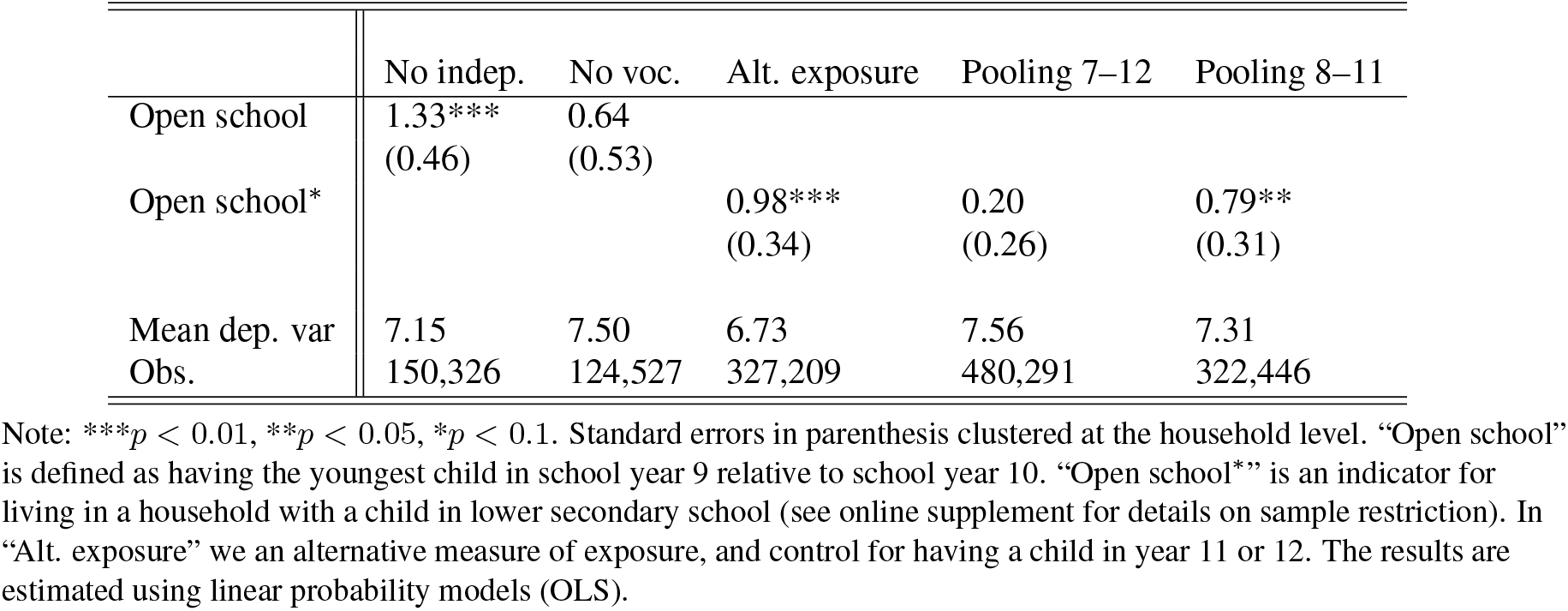
Robustness tests for parents. Outcome: Positive PCR tests per 1000.

**Table S5:** Robustness checks for teachers and teachers’ partners. Outcome: Positive PCR tests per 1000

|  | Teachers |  | Teachers' partners |  |
| --- | --- | --- | --- | --- |
|  | Excl. indep. schools | Excl. partner in healthcare | Excl. indep. schools | Excl. healthcare |
| Open school | 2.63***<br>(0.63) | 2.76***<br>(0.59) | 1.64**<br>(0.77) | 1.57**<br>(0.64) |
| Mean dep. var. | 2.96 | 2.83 | 5.10 | 2.78 |
| Obs. | 65,119 | 66,828 | 42,656 | 41,363 |

Note: \*\*\* $p < 0.01$ , \*\* $p < 0.05$ , \* $p < 0.1$ . Standard errors in parenthesis clustered the at the school level. "Open school" is defined as being a teacher or a teachers' partner at the lower secondary level. The results are estimated using linear probability models (OLS).

**Table S6:** Parents - Controlling for household size

|  | OLS |  | Logit |  |
| --- | --- | --- | --- | --- |
|  | PCR | Healthcare | PCR | Healthcare |
| Open school | 1.04**<br>(0.43) | -0.18<br>(0.26) | 1.17**<br>(0.07) | 0.93<br>(0.09) |
| Mean dep. var. | 6.37 | 2.74 |  |  |
| Obs. | 166,630 | 166,719 | 163,195 | 163,155 |

Note: \*\*\* $p < 0.01$ , \*\* $p < 0.05$ , \* $p < 0.1$ . Standard errors in parenthesis clustered the at the household level. “Open school” is defined as having the youngest child in school year 9 relative to school year 10. The results are estimated using linear probability models (OLS) and logistic regressions (Logit).

**Table S7:** Robustness check - different cutoff dates for the pre-period. Outcome: Positive PCR tests per 1000

|  | Parents |  | Teachers |  | Teachers' partners |  |
| --- | --- | --- | --- | --- | --- | --- |
|  |  |  | OLS (cases/1000) |  |  |  |
|  | March 25 | April 16 | March 25 | April 16 | March 25 | April 16 |
| Open school | 1.16***<br>(0.43) | 0.87**<br>(0.40) | 2.81***<br>(0.59) | 2.44***<br>(0.55) | 1.43**<br>(0.73) | 1.37**<br>(0.69) |
| Mean dep. var. | 6.54 | 5.74 | 2.96 | 2.66 | 5.32 | 4.59 |
| Obs. | 166,630 | 166,630 | 72,946 | 72,946 | 47,383 | 47,383 |
|  |  |  | Logit (odds ratios) |  |  |  |
|  | March 25 | April 16 | March 25 | April 16 | March 25 | April 16 |
| Open school | 1.18***<br>[1.04,1.33] | 1.16**<br>[1.01,1.32] | 2.01***<br>[1.52,2.67] | 1.96***<br>[1.46,2.64] | 1.27*<br>[0.99,1.64] | 1.30*<br>[0.99,1.71] |
| Obs. | 163,233 | 162,491 | 70,151 | 69,732 | 44,035 | 42,948 |

Note: \*\*\* $p < 0.01$ , \*\* $p < 0.05$ , \* $p < 0.1$ . Standard errors in parenthesis are clustered at the school level for teachers and their partners, at the household level for parents. "March 25" refers to moving the start of the investigation period to that date. Similarly, "April 16" moves the date to April 16. "Open school" is defined as having the youngest child in school year 9 relative to school year 10. The results are estimated using linear probability models (OLS) and logistic regressions (Logit).

**Table S8:**
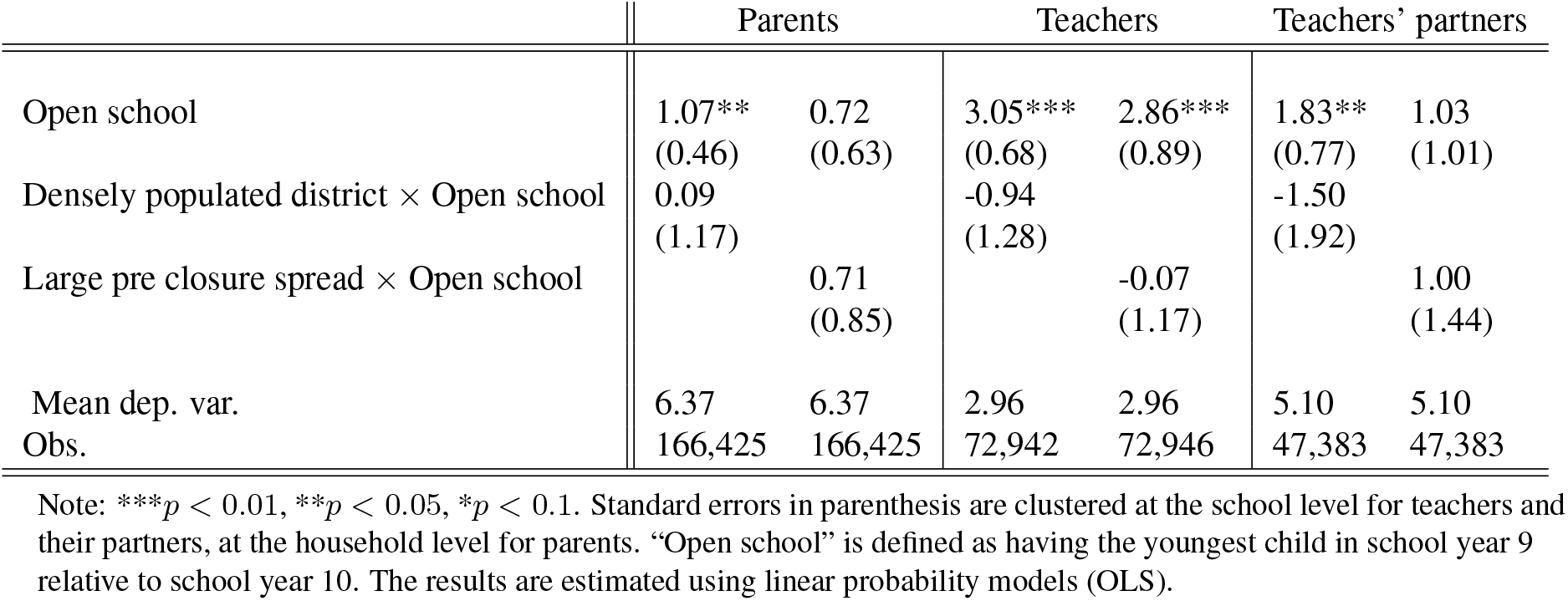
Heterogeneous treatment for teachers, teachers’ partners, and parents. Outcome: Positive PCR tests per 1000

|  | Parents |  | Teachers |  | Teachers' partners |  |
| --- | --- | --- | --- | --- | --- | --- |
| Open school | 1.07**<br>(0.46) | 0.72<br>(0.63) | 3.05***<br>(0.68) | 2.86***<br>(0.89) | 1.83**<br>(0.77) | 1.03<br>(1.01) |
| Densely populated district × Open school | 0.09<br>(1.17) |  | -0.94<br>(1.28) |  | -1.50<br>(1.92) |  |
| Large pre closure spread × Open school |  | 0.71<br>(0.85) |  | -0.07<br>(1.17) |  | 1.00<br>(1.44) |
| Mean dep. var. | 6.37 | 6.37 | 2.96 | 2.96 | 5.10 | 5.10 |
| Obs. | 166,425 | 166,425 | 72,942 | 72,946 | 47,383 | 47,383 |

Note: \*\*\* $p < 0.01$ , \*\* $p < 0.05$ , \* $p < 0.1$ . Standard errors in parenthesis are clustered at the school level for teachers and their partners, at the household level for parents. "Open school" is defined as having the youngest child in school year 9 relative to school year 10. The results are estimated using linear probability models (OLS).

**Table S9:** Students under age 18. Outcome: Positive PCR tests per 1000

|  | OLS (cases/1000) | Logit (odds ratios) |
| --- | --- | --- |
| School year 7 | -0.08<br>(0.13) | 0.86<br>[0.51,1.46] |
| School year 8 | -0.17<br>(0.13) | 0.70<br>[0.40,1.22] |
| School year 9 | -0.07<br>(0.13) | 0.89<br>[0.52,1.52] |
| Mean dep. var. | 0.53 |  |
| Obs. | 224,450 | 154,459 |

Note: \*\*\* $p < 0.01$ , \*\* $p < 0.05$ , \* $p < 0.1$ . Standard errors in parenthesis clustered at the school level. 95% confidence intervals in brackets are shown for the odds ratios. "School year ..." is in relation to school year 10 (reference category). The results are estimated using linear probability models (OLS) and logistic regressions (odds ratios).

**Table S10:** COVID-19 cases, patients and deaths by age group

| Age group | Cases | Cases<br>ex. health | Deaths | Deaths<br>ex. health | CFR (%) | CFR<br>ex. health | # patients<br>with diagnose | # patients<br>in hospital |
| --- | --- | --- | --- | --- | --- | --- | --- | --- |
| 0–6 | 152 | 152 | 1 | 1 | 0.66 | 0.66 | 132 | 67 |
| 7–16 | 457 | 457 | 0 | 0 | 0.00 | 0.00 | 230 | 94 |
| 17–19 | 614 | 611 | 0 | 0 | 0.00 | 0.00 | 230 | 84 |
| 20–29 | 5730 | 3204 | 7 | 7 | 0.12 | 0.22 | 2114 | 784 |
| 30–39 | 7396 | 3544 | 13 | 11 | 0.18 | 0.31 | 3185 | 1456 |
| 40–49 | 8586 | 4262 | 40 | 36 | 0.47 | 0.84 | 3997 | 1930 |
| 50–59 | 9978 | 5241 | 134 | 122 | 1.34 | 2.33 | 5275 | 3216 |
| 60–69 | 6463 | 4113 | 346 | 336 | 5.35 | 8.17 | 4666 | 3461 |
| 70–79 | 4792 | 4671 | 1112 | 1102 | 23.21 | 23.59 | 4756 | 3902 |
| 80– | 9314 | 9301 | 3749 | 3741 | 40.25 | 40.22 | 6050 | 5151 |
| Total | 53482 | 35556 | 5402 | 5356 | 10.10 | 15.06 | 30635 | 21045 |

Note: Test date until June 15, 2020. Deaths reported until July 25 for cases tested until June 15. "ex. health" means that healthcare and care workers are dropped (occupational codes 15, 22, 32, 53). CFR refers to the implied Case Fatality Rate.

**Table S11:** Occupations ranked by incidence of positive PCR-tests (lowest to highest incidence)

| Rank | Occupation title (SSYK3) | Cases/1000 | Occ. size |
| --- | --- | --- | --- |
| 1 | Specialists within environmental and health protection | 1.15 | 8690 |
| 2 | Mixed crop and animal breeders | 1.15 | 7807 |
| 3 | Animal breeders and keepers | 1.37 | 15315 |
| 4 | Museum curators and librarians and related professionals | 1.56 | 10931 |
| 5 | Architects and surveyors | 1.72 | 12218 |
| 6 | Mathematicians, actuaries and statisticians | 1.76 | 2267 |
| 7 | ICT architects, systems analysts and test managers | 1.93 | 127722 |
| 8 | Electronics and telecommunications installers and repairers | 1.96 | 10711 |
| 9 | Library and filing clerks | 1.98 | 3533 |
| 10 | Biologists, pharmacologists and specialists in agriculture and forestry | 2.00 | 7006 |
| 11 | Sheet and structural metal workers, moulders and welders, and related workers | 2.08 | 25959 |
| 12 | University and higher education teachers | 2.08 | 37457 |
| 13 | Designers | 2.11 | 16579 |
| 14 | Wood processing and papermaking plant operators | 2.19 | 15986 |
| 15 | Ships' deck crews and related workers | 2.22 | 1350 |
| 16 | Engineering professionals | 2.26 | 95496 |
| 17 | Market gardeners and crop growers | 2.27 | 23319 |
| 18 | Other service related workers | 2.31 | 3892 |
| 19 | Marketing and public relations professionals | 2.36 | 39061 |
| 20 | Metal processing and finishing plant operators | 2.37 | 16441 |
| 21 | Carpenters, bricklayers and construction workers | 2.44 | 106364 |
| 22 | Mobile plant operators | 2.46 | 35715 |
| 23 | Financial and accounting associate professionals | 2.49 | 57856 |
| 24 | Tax and related government associate professionals | 2.50 | 44063 |
| 25 | Recycling collectors | 2.55 | 8634 |
| 26 | Painters, Lacquerers, Chimney-sweepers and related trades workers | 2.57 | 26114 |
| 27 | Research and development managers | 2.58 | 6192 |
| 28 | Construction labourers | 2.59 | 6948 |
| 29 | Electrical equipment installers and repairers | 2.60 | 38434 |
| 30 | ICT operations and user support technicians | 2.61 | 44879 |
| 31 | Berry pickers and planters | 2.63 | 3045 |
| 32 | Physicists and chemists | 2.63 | 6462 |
| 33 | Forestry and related workers | 2.64 | 5690 |
| 34 | Creative and performing artists | 2.64 | 12491 |
| 35 | Accountants, financial analysts and fund managers | 2.65 | 49484 |
| 36 | Culinary associate professionals | 2.72 | 4046 |
| 37 | Broadcasting and audio-visual technicians | 2.76 | 4350 |
| 38 | Other stationary plant and machine operators | 2.76 | 7237 |
| 39 | Physical and engineering science technicians | 2.77 | 104914 |
| 40 | Information, communication and public relations managers | 2.78 | 4313 |
| 41 | Legal professionals | 2.80 | 22498 |
| 42 | Roofers, floor layers, plumbers and pipefitters | 2.90 | 35526 |
| 43 | Commissioned armed forces officers | 2.91 | 1032 |
| 44 | Postmen and postal facility workers | 2.91 | 15781 |
| 45 | Precision-instrument makers and handicraft workers | 2.94 | 4762 |
| 46 | Client information clerks | 2.96 | 60517 |
| 47 | Production managers in manufacturing | 2.98 | 16439 |
| 48 | Financial and insurance managers | 3.00 | 4995 |
| 49 | Printing trades workers | 3.05 | 7542 |
| 50 | Insurance advisers, sales and purchasing agents | 3.06 | 132527 |
| 51 | Machine operators, textile, fur and leather products | 3.07 | 5538 |
| 52 | Ship and aircraft controllers and technicians | 3.07 | 5535 |
| 53 | Real estate and head of administration manager | 3.08 | 3898 |
| 54 | Event seller and telemarketers | 3.09 | 9073 |
| 55 | Armed forces occupations, other ranks | 3.12 | 5448 |
| 56 | Information and communications technology service managers | 3.22 | 11185 |
| 57 | Blacksmiths, toolmakers and related trades workers | 3.22 | 49318 |
| 58 | Machinery mechanics and fitters | 3.23 | 59063 |
| 59 | Upper secondary school teachers | 3.24 | 32130 |
| 60 | Sports, leisure and wellness managers | 3.26 | 1533 |
| 61 | Shop staff | 3.27 | 194098 |
| 62 | Waiters and bartenders | 3.28 | 21963 |
| 63 | Dockers and ground personnel | 3.29 | 9415 |
| 64 | Production managers in construction and mining | 3.30 | 17554 |
| 65 | Finance managers | 3.36 | 17544 |
| 66 | Train operators and related workers | 3.38 | 5626 |
| 67 | Supply, logistics and transport managers | 3.39 | 11223 |
| 68 | Administrative and specialized secretaries | 3.43 | 17491 |
| 69 | Wood treaters, cabinet-makers and related trades workers | 3.46 | 11287 |
| 70 | Stores and transport clerks | 3.54 | 93586 |
| 71 | Administration and planning managers | 3.54 | 10438 |
| 72 | Sales and marketing managers | 3.57 | 30809 |
| 73 | Photographers, interior decorators and entertainers | 3.59 | 9198 |
| 74 | Vocational education teachers | 3.64 | 9888 |
| 75 | Authors, journalists and linguists | 3.67 | 16615 |
| 76 | Machine operators, rubber, plastic and paper products | 3.67 | 13342 |
| 77 | Business services agents | 3.69 | 32825 |
| 78 | Process control technicians | 3.69 | 18686 |
| 79 | Elected representatives | 3.74 | 1070 |
| 80 | Machine operators, food and related products | 3.77 | 14866 |
| 81 | Organisation analysts, policy administrators and human resource specialists | 3.79 | 112865 |
| 82 | Lower primary school teachers | 3.81 | 31992 |
| 83 | Assemblers | 3.88 | 55141 |
| 84 | Cashiers and related clerks | 3.97 | 11339 |
| 85 | Athletes, fitness instructors and recreational workers | 3.98 | 26376 |
| 86 | Manufacturing labourers | 4.05 | 10383 |
| 87 | Hotel and conference managers | 4.05 | 1483 |
| 88 | Construction and manufacturing supervisors | 4.05 | 24431 |
| 89 | Other services managers not elsewhere classified | 4.08 | 7103 |
| 90 | Mining and mineral processing plant operators | 4.14 | 7980 |
| 91 | Butchers, bakers and food processors | 4.14 | 8215 |
| 92 | Office assistants and other secretaries | 4.16 | 168407 |
| 93 | Architectural and engineering managers | 4.18 | 11232 |
| 94 | Croupiers, debt collectors and related workers | 4.22 | 2132 |
| 95 | Human resource managers | 4.27 | 8663 |
| 96 | Preschool managers | 4.28 | 4677 |
| 97 | Retail and wholesale trade managers | 4.46 | 10304 |
| 98 | Cooks and cold-buffet managers | 4.57 | 40728 |
| 99 | Heavy truck and bus drivers | 4.61 | 75333 |
| 100 | Administration and service managers not elsewhere classified | 4.70 | 23612 |
| 101 | Managing directors and chief executives | 4.71 | 20381 |
| 102 | Teaching professionals not elsewhere classified | 4.75 | 36664 |
| 103 | Upper primary school teachers | 4.82 | 29850 |
| 104 | Tailors, upholsterers and leather craftsmen | 4.85 | 3298 |
| 105 | Primary and secondary schools and adult education managers | 4.93 | 10557 |
| 106 | Building caretakers and related workers | 4.93 | 47028 |
| 107 | Cabin crew, guides and related workers | 4.96 | 8469 |
| 108 | Religious professionals and deacons | 4.98 | 3615 |
| 109 | Fast-food workers, food preparation assistants | 5.04 | 71276 |
| 110 | Newspaper distributors, janitors and other service workers | 5.12 | 41773 |
| 111 | Cleaners and helpers | 5.16 | 85416 |
| 112 | Other surveillance and security workers | 5.46 | 36460 |
| 113 | Washers, window cleaners and other cleaning workers | 5.49 | 7835 |
| 114 | Legislators and senior officials | 5.49 | 2915 |
| 115 | Hairdressers, beauty and body therapists | 5.53 | 21344 |
| 116 | Restaurant managers | 5.64 | 8332 |
| 117 | Driving instructors and other instructors | 5.8 | 7409 |
| 118 | Lower secondary school teachers | 5.83 | 37894 |
| 119 | Social work and counselling professionals | 6.69 | 46161 |
| 120 | Machine operators, chemical and pharmaceutical products | 6.9 | 5072 |
| 121 | Police officers | 8.08 | 16219 |
| 122 | Social work and religious associate professionals | 8.6 | 22084 |
| 123 | Education managers not elsewhere classified | 8.85 | 1243 |
| 124 | Car, van and motorcycle drivers | 9.03 | 19594 |

Note: Incidence (cases per 1000) of detected SARS-CoV-2 by 3-digit occupational codes (SSYK2012) until June 15, 2020. Ages 25–65, only occupations with at least 1000 employees reported. Healthcare occupations are excluded in the ranking. Teachers at different levels are identified using the Teacher Register and not by using SSYK codes 233 (compulsory school teachers) and 234 (upper secondary school teachers).

## Footnotes

1 1.47 cases per 1000 among partners and 2.81 cases per 1000 among teachers gives a SAR of 0.52. Bootstrapping with 2000 repetitions gives a non-parametric CI95 of 0.05–1.18.

## Notes

### Competing Interest Statement

The authors have declared no competing interest.

### Funding Statement

We are grateful for financial support from Handelsbankens forskningssiftelser.

### Author Declarations

This project was approved by the Swedish Ethical Approval Board on May 19, 2020 (decision number 2020-02323).

### Summary of Updates

Result section revised and expanded, in particular the robustness part; Figures 2 and 3 revised; Minor revisions on in tables 1 and 2, as well as some tables in the appendix due to a minor change in the population for a part of the study; Supplement updated.

